# Reproducible comparison and interpretation of machine learning classifiers to predict autism on the ABIDE multimodal dataset

**DOI:** 10.1101/2024.09.04.24313055

**Authors:** Yilan Dong, Dafnis Batalle, Maria Deprez

## Abstract

Autism is a neurodevelopmental condition affecting ∼1% of the population. Recently, machine learning models have been trained to classify participants with autism using their neuroimaging features, though the performance of these models varies in the literature. Differences in experimental setup hamper the direct comparison of different machine-learning approaches. In this paper, five of the most widely used and best-performing machine learning models in the field were trained to classify participants with autism and typically developing (TD) participants, using functional connectivity matrices, structural volumetric measures and phenotypic information from the Autism Brain Imaging Data Exchange (ABIDE) dataset. Their performance was compared under the same evaluation standard. The models implemented included: graph convolutional networks (GCN), edge-variational graph convolutional networks (EV-GCN), fully connected networks (FCN), auto-encoder followed by a fully connected network (AE-FCN) and support vector machine (SVM). Our results show that all models performed similarly, achieving a classification accuracy around 70%. Our results suggest that different inclusion criteria, data modalities and evaluation pipelines rather than different machine learning models may explain variations in accuracy in published literature. The highest accuracy in our framework was obtained by an ensemble of GCN models trained on combination of functional MRI and structural MRI features, reaching classification accuracy of 72.2% and AUC = 0.78 on the test set. The combined structural and functional modalities exhibited higher predictive ability compared to using single modality features alone. Ensemble methods were found to be helpful to improve the performance of the models. Furthermore, we also investigated the stability of features identified by the different machine learning models using the SmoothGrad interpretation method. The FCN model demonstrated the highest stability selecting relevant features contributing to model decision making. Code available at: https://github.com/YilanDong19/Machine-learning-with-ABIDE.

## 1 Introduction

Autism is a developmental condition characterized by deficits in social communication, social reciprocity, repetitive and stereotyped behaviors and interests (del Barrio 2004), and atypical responses to sensory stimuli (Green et al. 2013). Around 1% of the general population is diagnosed with autism (World Health Organization 2023), and symptoms such as early language delay usually appear when children are two to three years old (Landa 2008; Stefanatos 2008). Autism core symptoms and co-occurring conditions such as depression and learning disabilities (Stewart et al. 2006) often lead to serious challenges in the physical and mental well-being of the individuals with autism and their family. At present, the medical diagnosis of autism is largely based on behavioral observations and clinical interviews, and the underlying neural mechanisms of autism are still unclear (Yahata, Morimoto, and Hashimoto 2016). The insights provided by human neuroimaging studies may contribute to the development of autism biomarkers (Ecker, Bookheimer, and Murphy 2015).

In recent years, with the rapid development of magnetic resonance imaging (MRI) technology, we have gradually gained a deeper understanding of atypical brain function and structure linked to neurodevelopmental conditions such as autism. For instance, several structural MRI studies have reported differences in total brain volume (Courchesne 2002) and brain asymmetry (Postema et al. 2019) in participants with autism. Recent studies have also shown alterations in cortical thickness (CT) (Moradi et al. 2017) and subcortical volume (SV) (Katuwal et al. 2015).

Resting-state functional MRI (rs-fMRI) is widely used to describe long-range functional relationships between different brain regions in brain disorders (Du, Fu, and Calhoun 2018) and has been used to characterize atypical functional connectivity in autism. Widespread reductions in connectivity among children with autism have been reported, spanning unimodal, heteromodal, primary somatosensory, and limbic and paralimbic cortices, whereas those with autism had increased connectivity between a small set of nodes primarily in subcortical regions (di Martino et al. 2014). More recently, a pattern of hyperconnectivity in prefrontal and parietal cortices and hypoconnectivity in sensory-motor regions has been suggested to be consistent and reproducible across cohorts (Holiga et al. 2019).

However, neuroimaging studies investigating autism often report heterogeneous or even contradictory findings. For example, the volumetric differences between autism and typically developing participants (TD) reported by Aylward et al. (2002) were not replicated by Courchesne (2002). Differing results can be partially explained by biological heterogeneity within the population, variations in the imaging protocols, scanning parameters, and the scanner manufacturers, as well as variations in preprocessing and model evaluation methods. To date, no effective biomarker has been yet developed to enable a more objective diagnosis and stratification of autism, including the identification of sub-groups that may benefit from therapeutic interventions.

In addition to traditional statistical approaches, modern artificial intelligence techniques have recently been employed with the hope that they may help to uncover these differences. In the past ten years, classic machine learning techniques have been used to mine a wealth of information in structural MRI (sMRI) and rs-fMRI to conduct autism research. **Table 1** lists several experiments that were conducted using statistical and classical machine learning approaches. For instance, Anderson et al. (2011) used Pearson’s correlation coefficients to construct functional connectivity matrices from the time series of each pair of Regions of Interest (ROIs), then applied a two-tailed t-test to identify a subset of connections that were significantly different between the autistic and TD participants. They subsequently employed a linear classifier to classify individuals with autism from TD participants based on the selected connections, resulting in accuracy 79%. Ecker et al. (2010) performed one of the first studies in which autism was predicted based on structural brain measures. Segmented grey matter (GM) and white matter (WM) images comprised an input into a support vector machine (SVM) classifier, resulting in accuracy of 86%. Sabuncu and Konukoglu (2015) utilized the sMRI volumetric features from FreeSurfer software (grey matter volume, average thickness, etc.) to classify autistic participants, achieving 60% classification accuracy. Abraham et al. (2017) proposed a pipeline based on multi-subject dictionary learning (MSDL) atlas, tangent embedding and support vector classifier (SVC) with L2 regularization, achieving 67% prediction accuracy.

**Table 1.**
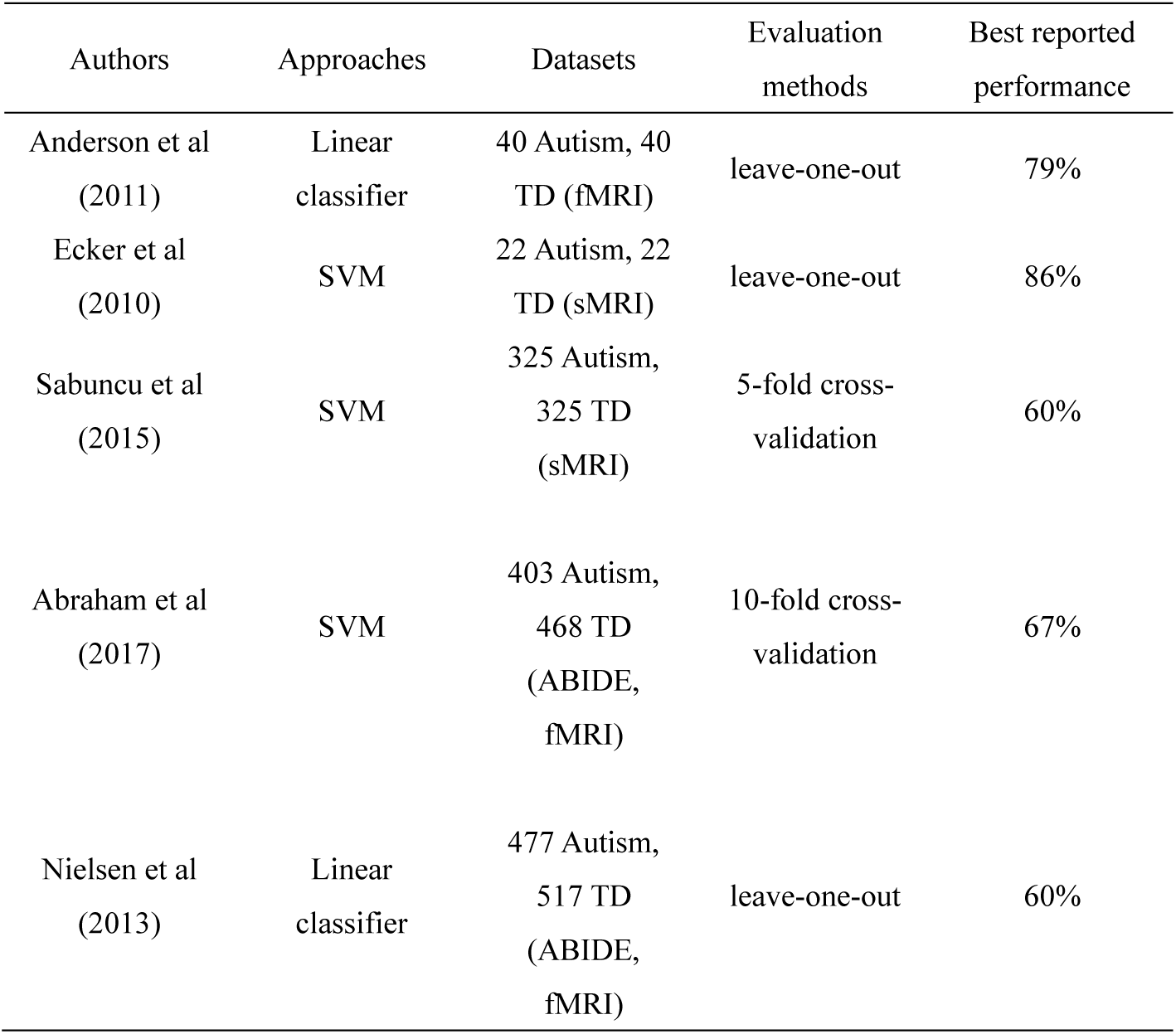
Summary of statistical and classical machine learning approaches predicting autism from MRI features.

However, other authors have found that the heterogeneity of data from multiple collection sites has a significant impact on the experimental results. For example, Nielsen et al. (2013) performed a similar experiment as Anderson et al. (2011) with a bigger sample size but obtained very different results. They utilised fMRI data from 964 participants (16 sites from ABIDE dataset) rather than 80 participants, which resulted in the classification accuracy decrease from 79% (Anderson et al. 2011) to 60% (Nielsen et al. 2013). Wolfers et al. (2019) summarized the results of 57 studies and observed a trend towards decreasing accuracy with increasing sample size in autism classification, suggesting that increasingly large sample sizes may result in low accuracies because of the intrinsic heterogeneity of autism, but may allow for the identification of more robust decision functions.

Deep learning models have achieved results comparable to human expert performance in many fields such as speech, natural language processing, and computer vision (Alasasfeh, Alomari, and Ibbini 2021; Ciregan, Meier, and Schmidhuber 2012). They are also widely applied in the medical imaging field, attempting to obtain more accurate experimental results than with classic machine learning approaches. **Table 2** lists several experiments conducted to predict autism using deep learning approaches. Heinsfeld et al. (2018) used deep neural networks with two auto-encoders, reaching 70% classification accuracy in distinguishing autistic and TD participants. With the supplement of structural MRI data, Rakić et al. (2020) achieved 85% accuracy by using auto-encoder followed by fully connected network classifier (AE-FCN) and Ensembles of Multiple Models and Architectures (EMMA), combining information from functional MRI and structural MRI. Parisot et al. (2018) developed a graph convolutional neural network (GCN) to incorporate phenotypic information. The best accuracy obtained by this model was 70.4%. Based on the original GCN model, Huang and Chung (2020) proposed an edge-variational graph convolutional neural network (EV-GCN) with reported classification accuracy of 81%.

**Table 2.**
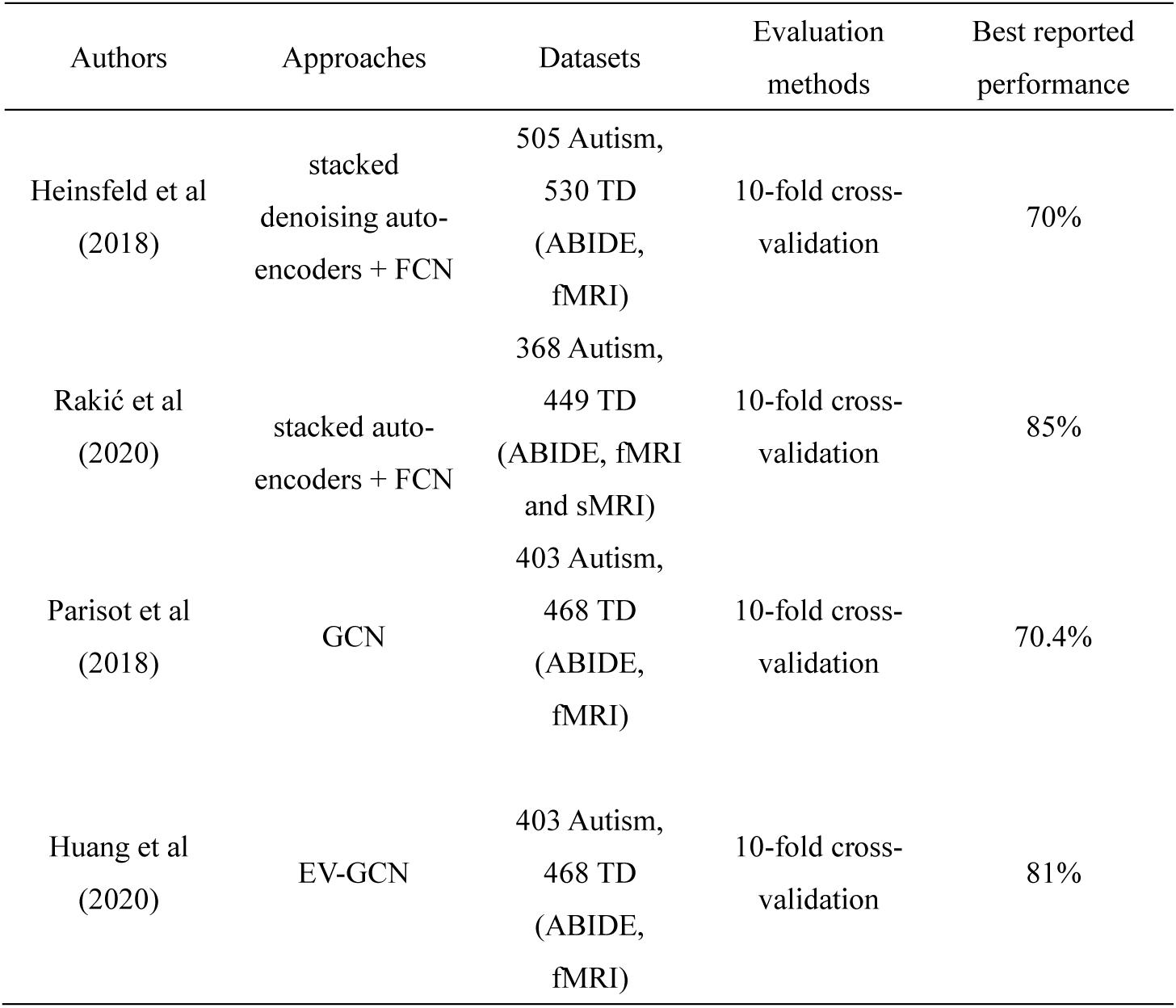
Summary of deep learning approaches predicting autism from MRI features.

The success of deep learning models stems from the combination of efficient learning algorithms and their large parameter spaces, which makes them considered complex black box models. This complexity renders machine decision-making opaque and significantly reduces the trust of doctors and patients in artificial intelligence (Singh, Sengupta, and Lakshminarayanan 2020). To overcome this weakness, intensive research on improving the interpretability of machine learning models has emerged, and a plethora of interpretation methods have been proposed to help researchers understand its inner workings mechanism (Salahuddin et al. 2022). For example, Layer-wise Relevance Propagation (LRP) as a gradient-based interpretation method (Montavon et al. 2017), was applied to a convolutional neural network (CNN) to reveal brain anatomical features associated with term and preterm birth (Grigorescu et al. 2019). Garg et al. (2020) utilized the Gradient-weighted Class Activation Mapping (Grad-CAM) (Selvaraju et al. 2016) and SmoothGrad (Smilkov et al. 2017) to picture the saliency images of the CNN model when classifying cancer histopathological images into malignant and benign categories.

In this study, we aimed to evaluate the performance of five previously proposed machine and deep learning models for the classification of autism under a standardised setting. We compared the performance of such models using a consistent repeatable framework, addressing inconsistencies in training datasets and evaluation frameworks reported in the literature. We proposed:

1. A standardized and comprehensive cross-validated evaluation framework, which fitted the models to the training set, tuned parameters on validation set and evaluated the performance on the test set. The whole fitting process was performed multiple times, while rotating the test set, so that the performance was evaluated on each sample exactly once. This framework both avoids any overfitting and gives a robust performance independent of the selection of the test set.
2. A selection of five of the most widely used or best-performing machine learning models from the existing literature, namely SVM (Bharadwaj, Prakash, and Kanagachidambaresan 2021), FCN (Rumelhart, Hinton, and Williams 2013), AE-FCN (Rakić et al. 2020), GCN (Parisot et al. 2018) and EV-GCN (Huang and Chung 2020), were used to classify autistic and TD participants using the ABIDE dataset. All classifiers were trained and evaluated using our standardized cross-validated evaluation framework to allow fair comparison of their performance.
3. To provide comprehensive evaluation, we trained our classifier with six different combination of features sets: (a) structural MRI (sMRI) features (b) sMRI + non-imaging features (c) functional MRI (fMRI) features (d) fMRI + non-imaging features (e) sMRI + fMRI features (f) sMRI + fMRI + non-imaging features
4. We compared the performance of the individual models, as well as an ensemble of the models trained on different subsets of the data.
5. We applied the SmoothGrad interpretation methods to GCN, FCN and AE-FCN to study model stability and understand what features contributed to model decision-making.

## 2 Methodology

### 2.1 ABIDE Dataset

The Autism Brain Imaging Data Exchange (ABIDE) database has aggregated brain structure and functional imaging data from multiple research institutes around the world to accelerate the research on the neural mechanisms of autism. Up to now, the ABIDE project has formed two large data sets: ABIDE I and ABIDE II (di Martino et al. 2014). In this work, we utilize extracted structural and functional features available for ABIDE I dataset (https://fcon_1000.projects.nitrc.org/indi/abide/abide_I.html).

#### 2.1.1 Participants

A total of 870 individuals (403 autistic and 467 TD participants) from 20 different collection sites were used in this study (**Table 3**). Our dataset was based on that used by Abraham et al. (2017), which comprises all ABIDE I participants, except for the ones with incomplete brain coverage and scanner artefacts. Additionally, we excluded one participant due to a FreeSurfer preprocessing failure.

**Table 3.** The 20 collection sites of ABIDE.

| Collection sites | N | Average age | Sex (male/female) | Autism/TD |
| --- | --- | --- | --- | --- |
| PITT | 50 | 18.50 | 43 / 7 | 24 / 26 |
| OLIN | 28 | 17.04 | 23 / 5 | 14 / 14 |
| OHSU | 25 | 10.81 | 25 / 0 | 12 / 13 |
| NYU | 172 | 15.33 | 136 / 36 | 74 / 98 |
| SBL | 26 | 33.77 | 26 / 0 | 12 / 14 |
| SDSU | 27 | 14.36 | 21 / 6 | 8 / 19 |
| STANDFORD | 25 | 9.99 | 18 / 7 | 12 / 13 |
| TRINITY | 44 | 17.03 | 44 / 0 | 19 / 25 |
| UCLA_2 | 21 | 12.47 | 19 / 2 | 11 / 10 |
| UM_1 | 86 | 13.77 | 61 / 25 | 34 / 52 |
| UM_2 | 34 | 16.01 | 32 / 2 | 13 / 21 |
| USM | 67 | 22.59 | 67 / 0 | 43 / 24 |
| YALE | 41 | 13.31 | 25 / 16 | 22 / 19 |
| CALTECH | 15 | 26.79 | 10 / 5 | 5 / 10 |
| CMU | 11 | 26.82 | 7 / 4 | 6 / 5 |
| KKI | 33 | 10.31 | 24 / 9 | 12 / 21 |
| LEUVEN_1 | 28 | 22.43 | 28 / 0 | 14 / 14 |
| LEUVEN_2 | 28 | 14.17 | 21 / 7 | 12 / 16 |
| MAX_MUN | 45 | 26.49 | 41 / 4 | 19 / 26 |
| UCLA_1 | 64 | 13.35 | 55 / 9 | 37 / 27 |

#### 2.1.2 Non-imaging features

Each sample used in this paper had several phenotypic and fMRI image quality measures, including age, gender, collection site, full scale IQ (FIQ), the number of timepoints with motion outliers (NUM), percentage of timepoints with motion outliers (PEC), and quality control anatomical rate (RAT). We used the collection site information in the GCN model (Sec. 2.3.4) to generate the connections between samples in the population graph.

#### 2.1.3 Functional MRI features

We downloaded the preprocessed fMRI data from ABIDE website directly (http://preprocessed-connectomes-project.org/abide/download.html). Briefly, the preprocessing steps include skull striping, slice timing correction, motion correction, global mean intensity normalization, nuisance signal regression, and band-pass filtering (0.01-0.1Hz). The fMRI BOLD timeseries were averaged for each brain region of the Cameron Craddock’s 200 ROI (CC200) atlas (Craddock et al. 2012) and Pearson’s correlation coefficients were calculated for each pair of ROIs to construct functional connectivity matrices for each participant.

#### 2.1.4 Structural MRI features

All the raw sMRI data were pre-processed by FreeSurfer software with *‘recon-all’* command, which includes intensity normalization, skull stripping, registration of the volumes to a common space, segmentation, cortical, white matter and subcortical parcellation.

Desikan-Killiany cortical atlas with 68 regions was selected as the atlas for cortical parcellation (Desikan et al. 2006)), each region was assigned 9 features: number of vertices, surface area, gray matter volume, average thickness, thickness standard deviation (SD), integrated rectified mean curvature, integrated rectified gaussian curvature, folding index and intrinsic curvature index. We also used metrics from 115 non-cortical regions (white matter, ventricles and sub-cortical regions) (Fischl et al. 2002; Salat et al. 2009). Each non-cortical region was assigned 7 features: number of voxels, volume, normalized intensity mean, normalized intensity SD, normalized intensity minimum, normalized intensity maximum, and normalized intensity range. The selection criteria of structural features are introduced in Sec. 2.2.2. In **Table 4**, statistical outputs evaluated are summarized. Among these, we selected statistical outputs from the cortical parcellation, subcortical and white matter as the structural features.

**Table 4.**
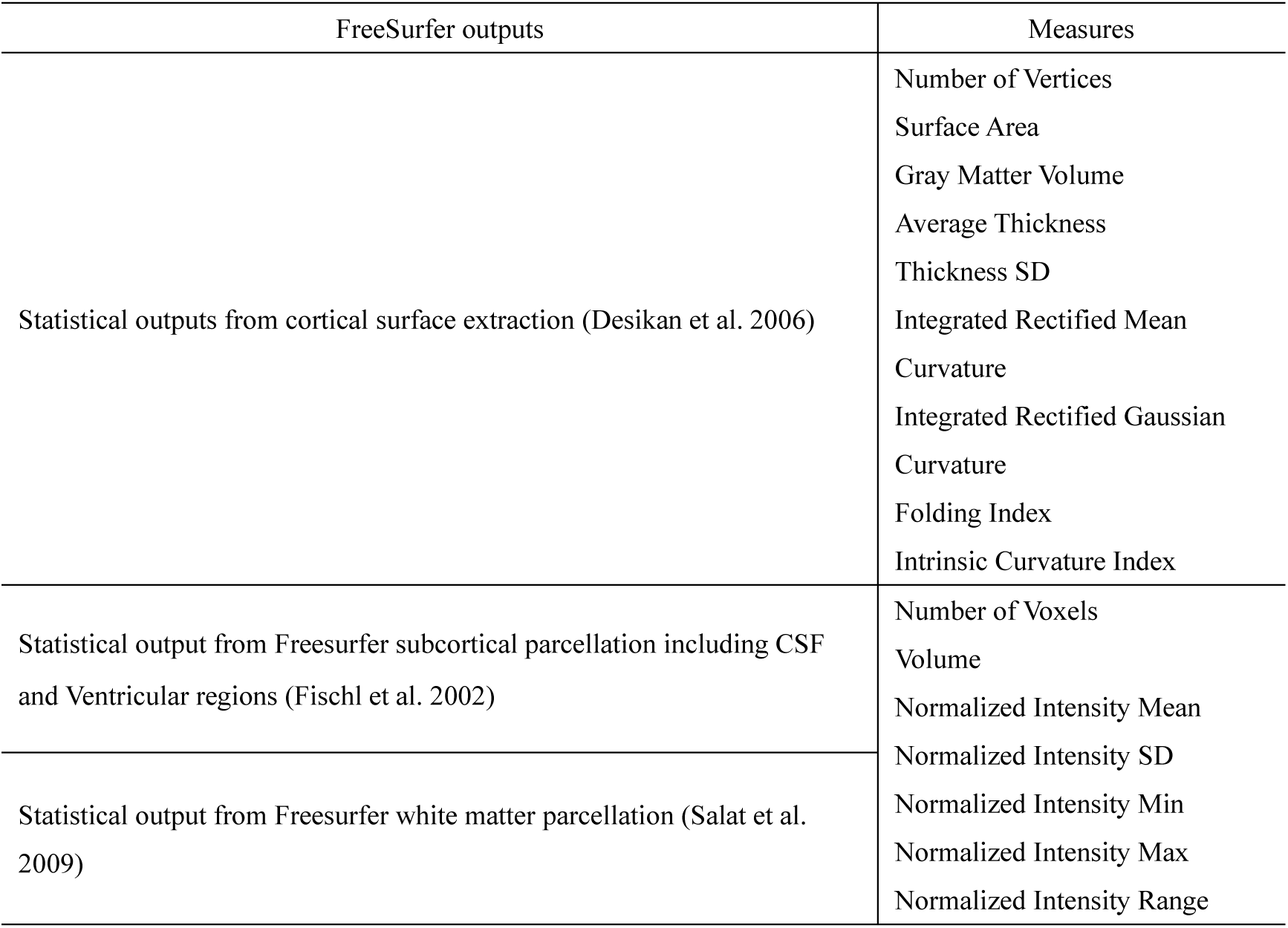
The FreeSurfer statistical outputs evaluated.

### 2.2 Input Features

Due to a large number of features, especially in functional connectivity matrices, feature selection is a necessary step before features are input to the model, ensuring that the model is not overfitted and generalizes to new data well. Inputting all connectivity features into the models would lead to overfitting, decreasing model performance on the test set. We performed feature selection on fMRI and sMRI features separately to create input for the fMRI only and sMRI only models. For the joint model, we concatenated the selected sMRI and fMRI features.

#### 2.2.1 fMRI Features

Due to the symmetry of the functional connectivity matrices, we first extracted the upper triangle of the functional connectivity matrix for each sample. the selected features were standardized to remove the mean and scale to unit variance, making sure all the features had similar distributions. We then performed recursive feature elimination using a ridge classifier. This approach has been previously shown to yield better results than alternative dimensionality reduction methods like auto- encoders and principal component analysis (PCA) (Parisot et al. 2018).

To determine an appropriate number of functional features for training of the machine learning models, we evaluated performance of baseline kernel SVM classifier (gamma=’scale’) using the grid search of different numbers of selected input features (1000, 2000,…, 19900) produced by recursive feature elimination. We found that selecting 4000 features with a regularization parameter of C =1 resulted in optimal cross-validated accuracy in prediction of autism and TD classes. Therefore, we select 4000 functional connectivity features, flattened into a 1-D array, as the fMRI input for the machine learning models.

#### 2.2.2 sMRI Features

We selected statistical outputs from cortical surface extraction, and subcortical and white matter parcellations as structural features (**Table 4**). From the three cortical parcellation atlases provided by FreeSurfer, we chose Desikan-Killiany Atlas, considering it is widely used in the literature (Mizuno et al. 2019; Zabihi et al. 2019).

To create the sMRI input features, we concatenated the cortical surface extraction output features for the Desikan-Killiany atlas, and subcortical and white matter parcellation outputs. This resulted in 1417 structural features flattened into a 1-D array. Due to the variation in feature distribution, standardization is applied to all the features to set the mean to zero and scale to unit variance. We then applied the SVM classifier using the grid search of different numbers of selected input features (100, 200, …, 1400, 1417) produced by RFE. We selected 800 structural features flattened into a 1-D array as the optimal number of sMRI features to input in further machine learning models.

#### 2.2.3 Non-imaging features

Considering each participant has only 6 non-imaging measures: age, gender, FIQ, NUM, PEC, RAT, we simply concatenated them together without employing feature selection when preparing the input features for the following three scenarios: (a) sMRI + non-imaging features (b) fMRI + non-imaging features (c) sMRI + fMRI + non-imaging features.

### 2.3 Machine learning models

In this paper, we investigated five machine learning models to train classifiers to predict Autism and TD classes: support vector machine (SVM) (Bharadwaj et al. 2021), fully connected network (FCN) (Rumelhart et al. 2013), auto-encoder followed by the fully connected network (AE-FCN) (Rakić et al. 2020), graph convolutional network (GCN) (Parisot et al. 2018) and edge-variational graph convolutional network (EV-GCN) (Huang and Chung 2020).

#### 2.3.1 Kernel SVM

We chose a Kernel SVM model with Gaussian kernel (‘rbf’) to create a benchmark classifier to compare with neural network-based models, as shown in **Fig. 1**. It is a fast and flexible non-linear classifier with tunable kernel size and regularization (Bharadwaj et al. 2021).

**Fig. 1.**
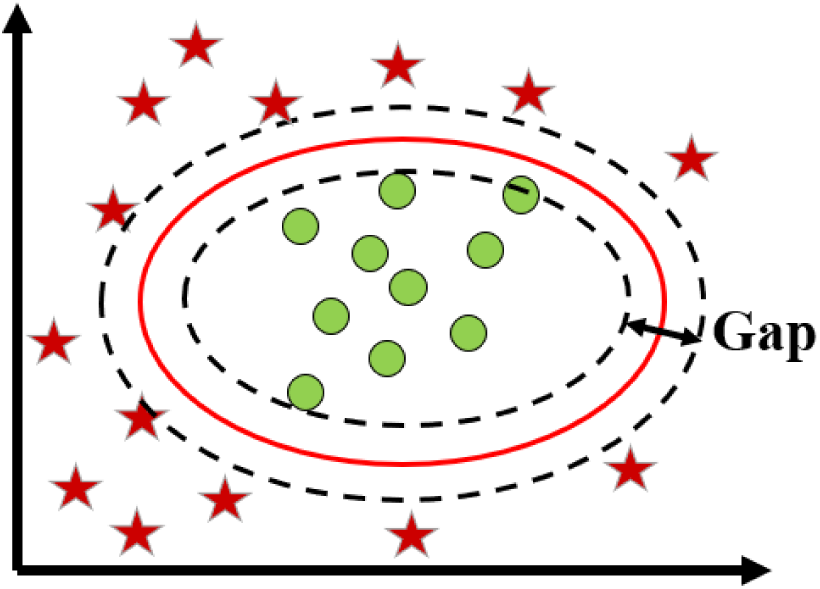
A support vector machine (SVM) for two-group classification problems.

The kernel SVM is implemented in scikit-learn. We select the Gaussian kernel with kernel size set to ‘scale’. The regularization parameter ‘C’ is selected using the grid search algorithm through cross-validation, and is adapted separately for each fold.

#### 2.3.2 FCN

The first deep neural network applied was a fully connected neural network (FCN) (Rumelhart et al. 2013). Its architecture is shown in **Fig. 2**

**Fig. 2.**
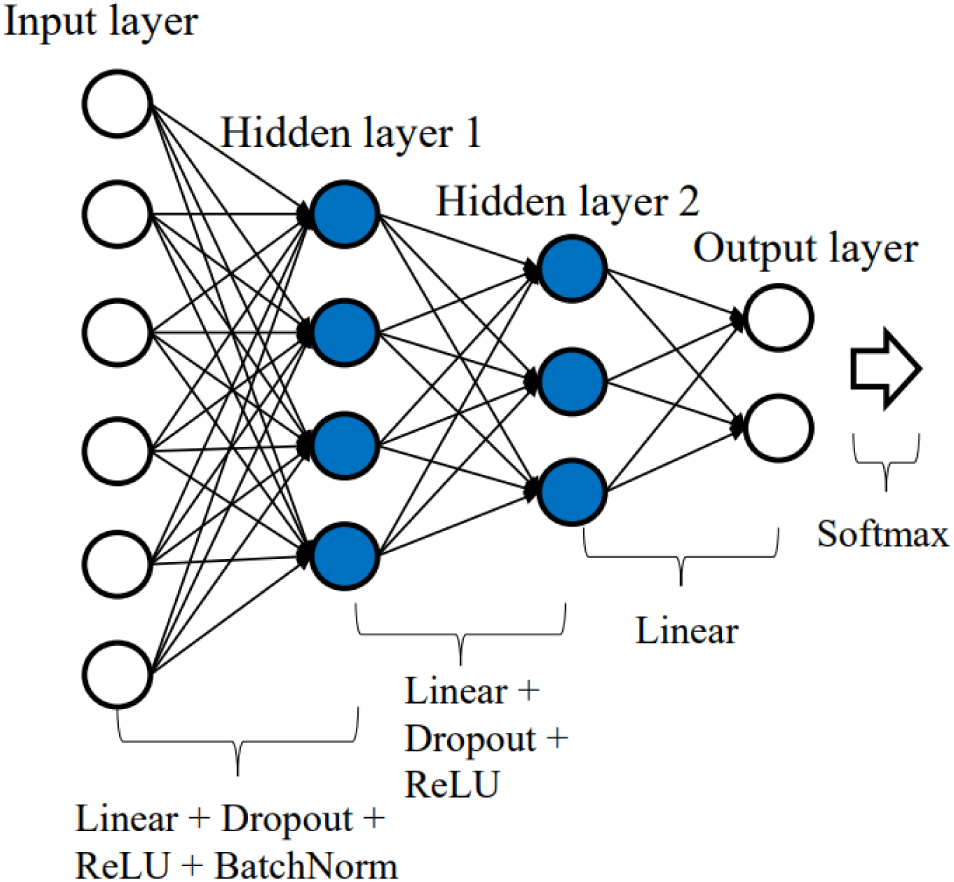
The structure of FCN. Linear layers are fully connected with each other. The white arrow on the right is the softmax layer, its output will be used to calculate cross entropy loss with labels. The number of nodes in each layer: 5000 (input), 500 (hidden 1), 30 (hidden 2), 2 (output).

The network consisted of three linear (fully connected) layers. To prevent this model from overfitting, the dropout layers (dropout rate = 0.5) were added after the first and second linear layers.

#### 2.3.3 Auto-encoder + FCN

Rakić et al. (2020) applied an auto-encoder followed by the fully connected network (AE-FCN) to the autism classification problem. This architecture aims to reduce the dimension of the input features by creating a bottleneck. The performance was further improved by using Ensembles of Multiple Models and Architectures (EMMA) (Kamnitsas et al. 2017).

We have implemented AE-FCN based on the description provided by Rakić et al. (2020). We optimized the parameters of the network ourselves as they were not detailed in the original study. The structure of our optimized model is shown in **Fig. 3**.

**Fig. 3.**
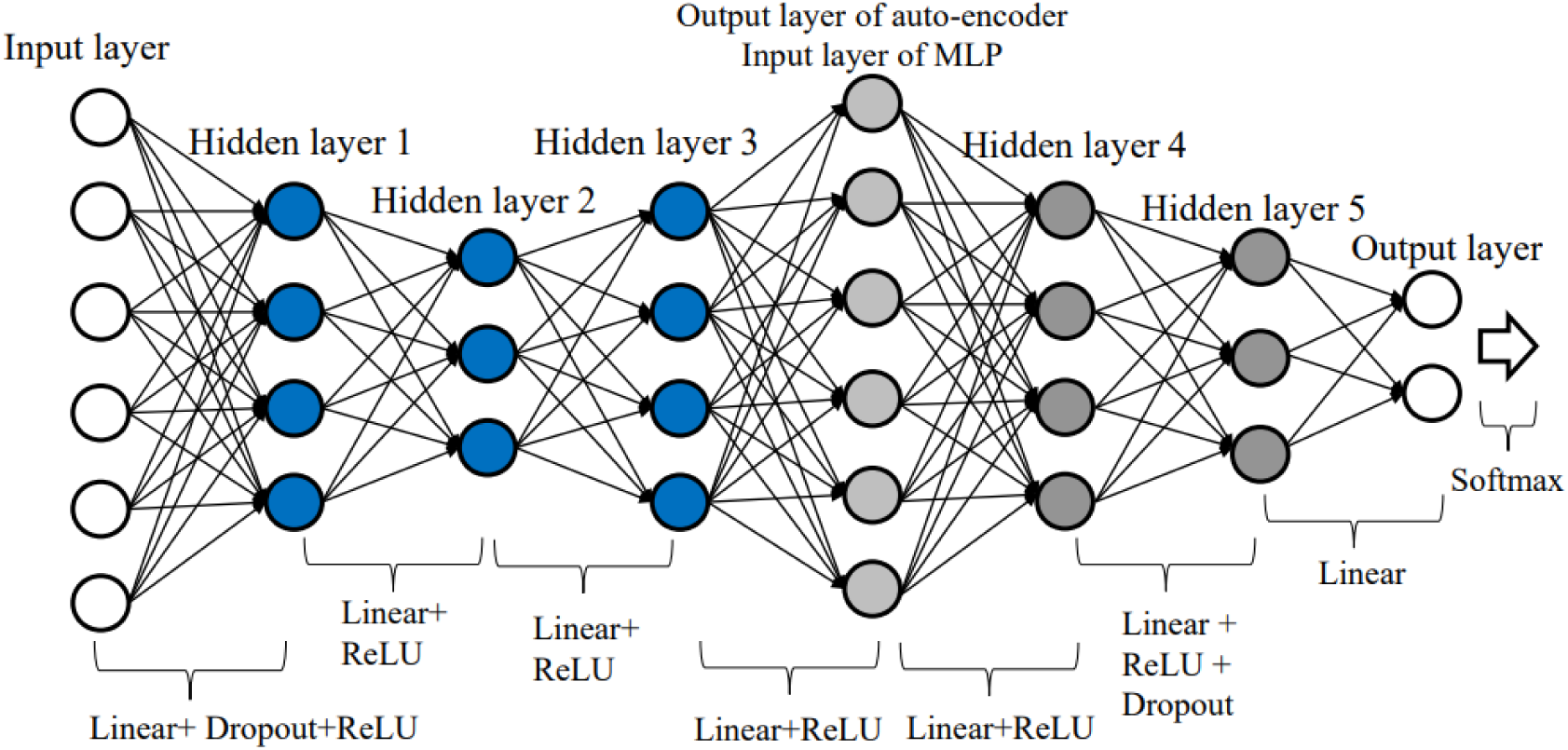
The structure of AE-FCN. The number of nodes in hidden layers 1 and 3 are the same, and hidden layers 2 and 5 have the same number of nodes. Mean squared error (MSE) loss is applied to train the auto-encoder to produce output similar to input, while cross-entropy loss is applied to give the correct prediction for each individual. The total loss is equal to MSE loss plus cross-entropy loss. The number of nodes in each layer: 5000 (input), 300 (hidden 1), 150 (hidden 2), 300 (hidden 3), 5000 (output layer of auto-encoder and input layer of FCN), 300 (hidden 4), 16 (hidden 5), 2 (output).

#### 2.3.4 GCN

Graph convolutional network (GCN) was proposed for autism classification by Parisot et al. (2018). This network allows to process the samples based on their similarity, which can be calculated from imaging, phenotypic and acquisition related information, to reduce the dimensionality of the feature space.

The structure of the GCN model used in this paper is shown in **Fig. 4**. First, a population graph that reflects the similarity of individual samples needs to be calculated. The goal is to leverage the complementary non-imaging information to calculate similarities between participants to create a graph structure and thus exploit the power of graph convolutions (Parisot et al. 2018). In our implementation, the subject-to-subject similarity, which are used as the weights of the edges of the graph, were calculated to indicate whether the participants were imaged in the same (weight=1) or different (weight=0) collection site. We experimented with other designs of the population graph that included other phenotypic and image quality information; however, we did not find any benefit in performance compared to only including site information.

**Fig. 4.**
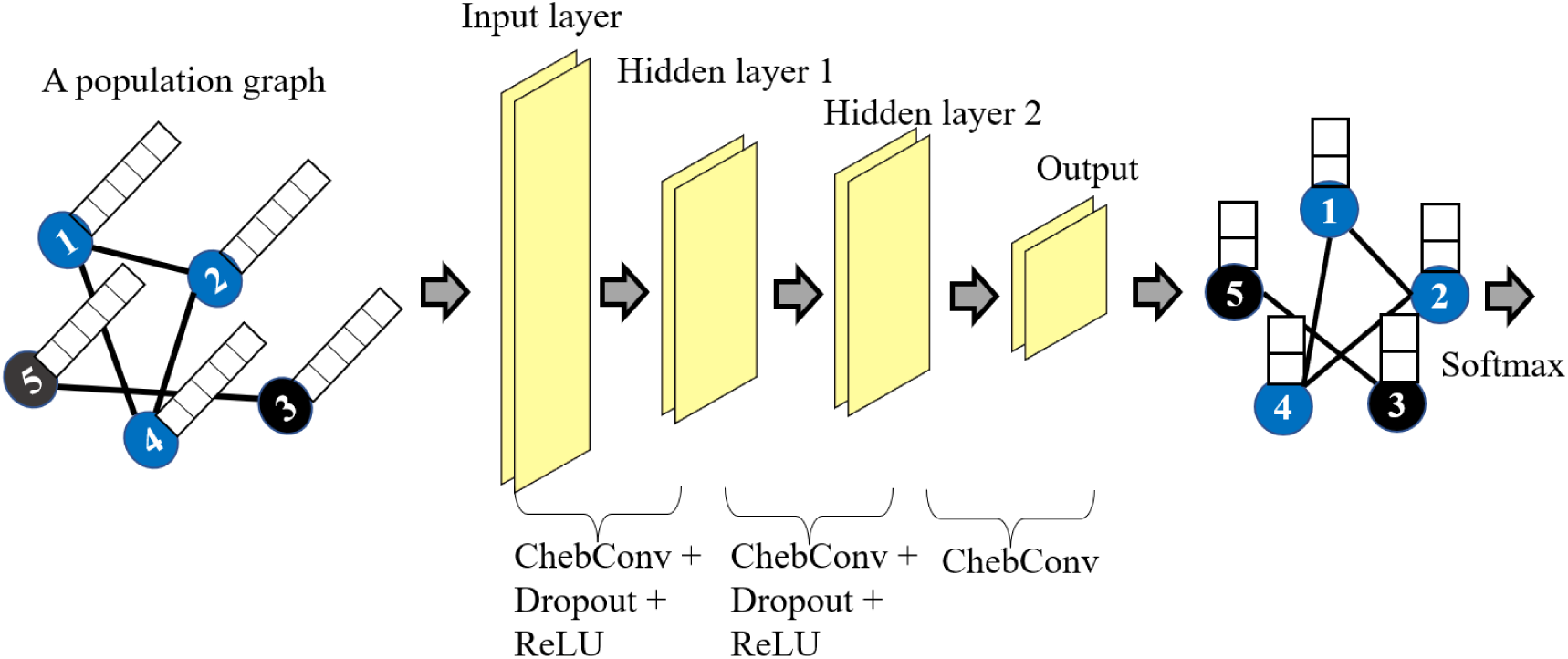
The structure of 3 layers GCN model. In the population graph, only the nodes with the same color have a connection between them (black line). Features of each sample are the white squares on the node. Cross entropy loss is applied at the end. The feature channels in each layer: 5000 (input), 128 (hidden 1), 128 (hidden 2), 2 (output).

#### 2.3.5 EV-GCN

Edge-variational graph convolutional network (Huang and Chung 2020) has been shown to significantly outperform GCN (Parisot et al. 2018) see **Table 2**. **Fig. 5** presents the structure of EV- GCN. It consisted of a pairwise association encoder (PAE), edge dropout layer (ED), four Chebyshev graph convolution layers, and one fusion block followed by two fully connected layers. The pairwise association encoder (PAE) generates an adaptive population graph, therefore connections change during the training process. The fusion block fuses the hidden features in each depth to alleviate the over-smoothing problem in deep GCN models.

**Fig. 5.**
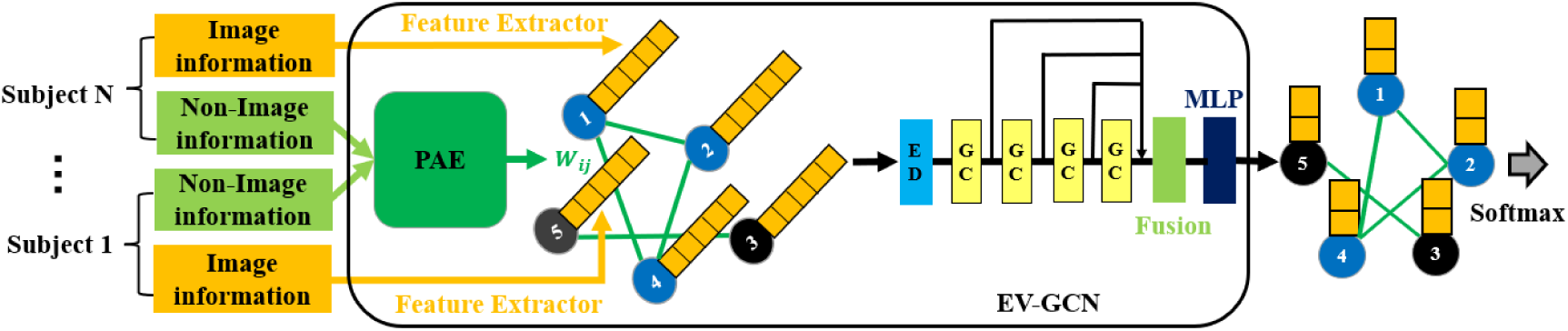
The structure of EV-GCN (Huang and Chung 2020). The connections in the Adaptive Population Graph are constructed by PAE block with only non-images (phenotypic) information, and they will be changed in the training process by the backpropagation of the cross-entropy loss function. ED: edge dropout layer. GC: graph convolution layer. The parameters in each GC layers are: 5000(input), 16 (hidden 1), 16 (hidden 2), 16 (hidden 3), 16 (output). Fusion block: concatenate the 4 outputs from previous GC layers. MLP: multilayer perceptron, with parameters: 64(input, 64 = 16*4), 256 (hidden 1), 2 (output).

We have adapted publicly available EV-GCN code (https://github.com/SamitHuang/EV_GCN) and applied it to the ABIDE dataset in our evaluation framework (see Sec. 2.5). EV-GCN constructs the adaptive population graph by inputting ‘gender’ and ‘site’ phenotypic information to PAE. In contrast to Huang and Chung’s approach of taking only the upper triangular of the population graph as the graph input, we take a different approach in EV-GCN by inputting the entire population graph.

### 2.4 Ensemble methods

The ensemble methods combine multiple machine learning models to improve classification performance. We investigated two types of ensembles: max voting and Ensembles of Multiple Models and Architectures (EMMA).

In max voting, multiple classification models are used to make predictions for each data point, and the prediction of each model is considered a “vote.” The final predictions are the labels that obtained the majority of the votes (Thomas G. Dietterich 2000). We applied max voting to aggregate responses from the five models of the same architecture trained during cross-validation (See Sec. 2.5) The purpose of EMMA is to obtain robust performance by aggregating the predictions from models with different architectures (Kamnitsas et al. 2017). In our experiments we used EMMA to combine the outputs of all five models considered in this paper (SVM, FCN, AE-FCN, GCN, EV- GCN) by majority voting.

In addition to the predictions using ensemble methods, we also measured the performance of the individual models, denoted as ‘no ensemble’ in our results.

### 2.5 Evaluation pipeline

The evaluation pipeline can have important effects on the measured performance of the machine learning models. We propose a robust evaluation framework based on two principles. Firstly, it is important to have a training set for model fitting, a validation set for hyper-parameter tuning and a test set for the performance measurement to avoid model overfitting and consequently artificially increasing the performance that cannot be repeated on unseen datasets. Secondly, the performance should be calculated using all samples in the dataset to avoid performance variation caused by variability in sample set selection on which the method is evaluated. We propose two different evaluation frameworks, one for the individual machine learning models, and the second one for the ensembles. These frameworks are illustrated in **Fig. 6**.

**Fig. 6.**
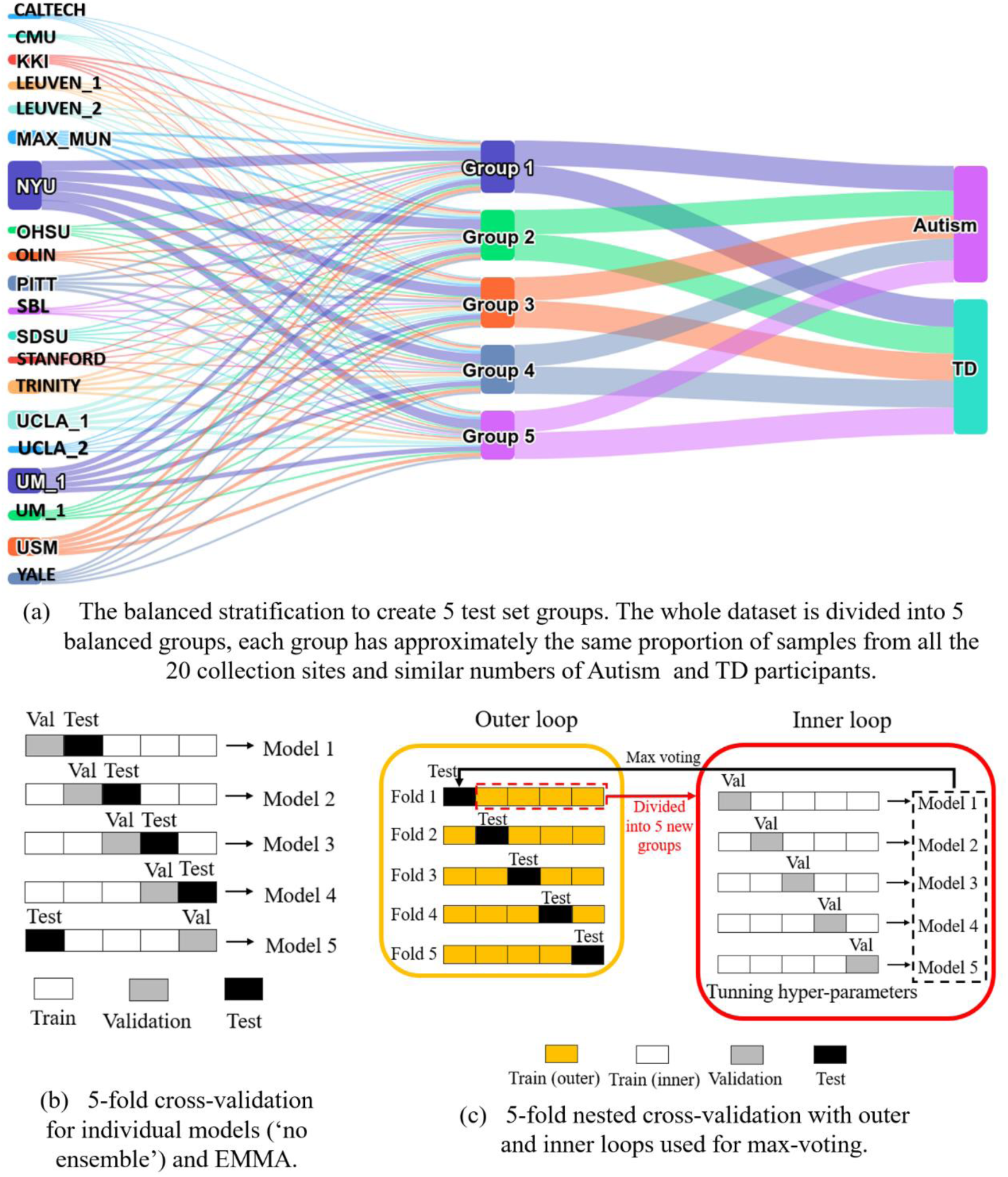
Diagrams of our evaluation pipelines. We used the same 5 test set groups (a) for all three types of models (‘no ensemble’, max-voting and Emma). The cross-validation approach (b) was applied to individual models and EMMA, while nested cross-validation (c) is designed to enable the max voting of five models of the same type.

#### 2.5.1 Creation of test sets

Our cross-validation approach started by splitting the whole dataset into 5 groups (**Fig. 6a**). Each of these groups will act as the test set exactly once, while we train the models using the remaining 4 groups. Therefore, the training process will be performed 5 times. This way performance can be evaluated on all the samples, while also keeping the model training and tuning completely independent of the test set.

While this is commonly done randomly, we instead opt for a fixed setup where we stratify the groups to have the same proportions of samples of Autism and TD labels and collection sites (**Fig. 6a**). This robust set up ensures consistency in performance measurements across our experiments. We have opted for 5 folds, because it provides a good compromise between robustness and the number of models that need to be trained. Additionally, it would be difficult to properly stratify the data into more folds according to all our variables due to the reduced sample size available from some collection sites.

#### 2.5.2 Cross-validation

For evaluation of each of the five individual models listed in Sec. 2.3 (‘no ensemble’), we used a 5- fold cross-validation approach presented in (**Fig. 6b**). In this set-up the model was tuned five times. In each fold, we have one test set to measure performance, one validation set that was used to select the optimal hyper-parameters, and the remaining three groups were used for training. The model was fitted to the training data multiple times with different hyper-parameter values, and hyper- parameters were selected according to the highest performance on the validation set in each fold. After the hyper-parameters had been tuned, the performance of the selected model in each fold was evaluated on the test set. The final performance was calculated by averaging performance on the test sets over all five folds. We calculated the accuracy and area under ROC curve (AUC) for each machine learning model, and applied two-sample paired T-test to compare the performance of different models, feature sets and ensemble methods.

For the kernel support vector classifier, we tuned the kernel size and the regularization parameter ’C’. For the deep learning models, we selected the number of iterations in each fold according to the performance on the validation set.

To predict the results using EMMA algorithm, we aggregated the predictions from SVM, FCN, AE- FCN, GCN and EV-GCN from all five folds, and obtained the final prediction by majority voting.

#### 2.5.3 Nested Cross-validation

In the case of max voting ensemble technique, we applied a nested cross-validation approach (Cawley and Talbot 2010) presented in **Fig. 6c**. In each outer fold, we selected the test set as detailed in Sec. 2.5.1 and **Fig. 6a**, and the remaining 4 groups were further divided into 5 new groups in the inner loop (using the same stratification strategy as before) for training and tuning of the model using 5-fold cross-validation. This resulted in training 25 different models for each architecture. We selected the hyper-parameters (kernel size and regularization for SVM, iteration number for DL models) based on the best average performance during 5-fold cross-validation, separately within each outer fold. Once the hyper-parameters were chosen for each outer fold, the 5 models trained during the inner cross-validation loop with the selected hyper-parameters voted on the predicted labels on the test set.

### 2.6 The SmoothGrad interpretation method

The gradients of machine learning models play an important role in interpretability analysis. Analyzing the gradients of the model’s output with respect to input features allows estimation of the relative importance of different features (Linardatos, Papastefanopoulos, and Kotsiantis 2021). Several gradient-based interpretation methods, including LRP, SmoothGrad, and Grad-CAM (Montavon et al. 2017; Selvaraju et al. 2016; Smilkov et al. 2017), have been proposed previously. However, considering the application conditions of each method, we ultimately opted for SmoothGrad as the interpretability method in this paper for investigating model stability and explaining the models’ decision, given that SmoothGrad is applicable to various architectures of deep learning models, and its mechanism is straightforward, relying solely on the gradient of the model. Applying SmoothGrad to the machine learning model yields a saliency map that indicates the importance of each input feature in the model’s decision-making process.

Due to the sensitivity of raw gradients to minor perturbations (noise) in the input, which results in unstable interpretation outcomes, the visualization of raw gradients extracted from machine learning models typically exhibits a high degree of noise (Smilkov et al. 2017). To mitigate this sensitivity to noise, SmoothGrad introduces random noise into the model’s inputs, creating multiple noisy inputs for each participant. The gradients from these noisy inputs are averaged to reduce the sensitivity to noise, yielding a smoother and more stable interpretation result (Smilkov et al. 2017). In this paper, we generated 10 noisy inputs for each participant to compute the average SmoothGrad saliency maps, resulting in stable interpretation results.

To obtain reliable important features from SmoothGrad, it is essential to consider the stability of the machine learning model. We expect the stable model to maintain consistent performance or output results when faced with data variations and assign high saliency values to features consistently. In the 5-fold nested cross-validation pipeline (**Fig. 6c**), we have 25 models (with the same model architecture) trained on different training sets, a stable machine learning model should provide us with consistent saliency maps for all these models. These consistent maps indicate the focus of the model on the same group of important features that contribute to the model’s decisions, even across varying training sets. In order to compare model stability, we have developed a measure of saliency stability akin to a “signal-to-noise ratio” (SNR) (**Fig. 7**) of saliency maps for each machine learning model (to compare between GCN, FCN and AE-FCN).

**Fig. 7.**
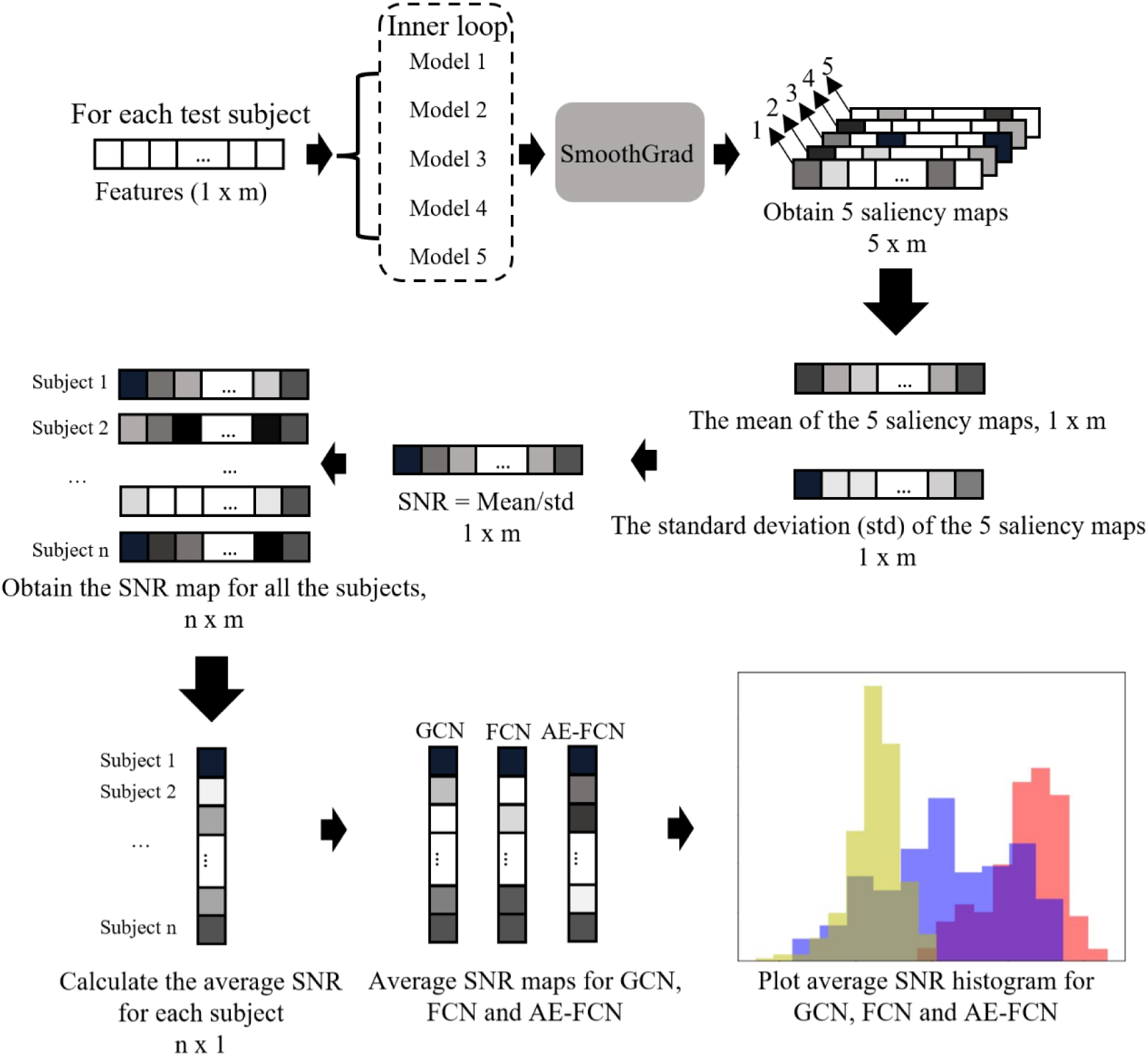
Stability assessment diagram of machine learning models.

The detailed model stability evaluation pipeline is shown in **Fig. 7**. For each test participant, we employ SmoothGrad to generate 5 saliency maps from its corresponding 5 models, and subsequently compute the feature-wise mean, standard deviation (std) and SNR from these 5 saliency maps. We then calculate the average SNR value (mean/std) for each participant and compare the average SNR histogram across different machine learning models. To minimize the bias caused by the model initialization, we repeat the 5-fold nested cross-validation pipeline 3 times for GCN, FCN and AE- FCN to calculate the average SNR maps. The most important features contributing to the model’s decision were extracted from the saliency maps of the most stable model.

In the stability experiment, we excluded the SVM and EV-GCN models for the following reasons: SmoothGrad cannot be applied to the SVM model; EV-GCN shares the same model type with GCN but with lower performance.

## 3 Results

We have performed a comprehensive evaluation of machine learning models performances using consistent evaluation framework presented in section 2.5. In section 3.1, we aimed to compare performance of:

- Five machine learning models detailed in section 2.3 (SVM, FCN, AE-FCN, GCN, EV- GCN)
- Three label fusion strategies detailed in section 2.4 (‘no ensemble’, max-voting, EMMA)
- Six different feature sets (a) structural MRI (sMRI) features (b) sMRI + non-imaging features (c) functional MRI (fMRI) features (d) fMRI + non-imaging features (e) sMRI + fMRI features (f) sMRI + fMRI + non-imaging features

In section 3.2, we assessed the stability (“SNR” values) of different machine learning models using SmoothGrad. Upon determining the most stable machine learning model, we extracted the most important features from the model’s saliency map in section 3.3.

### 3.1 Classification task

The performances of all models in terms of prediction accuracy and area under ROC curve (AUC) are presented in **Table 5**. We present average performance on validation set and test set. Note that the performance is aggregated from all cross-validation folds, therefore each sample contributed to the final performance exactly once. The model performances are further visualised in **Fig. 8**.

**Fig. 8.**
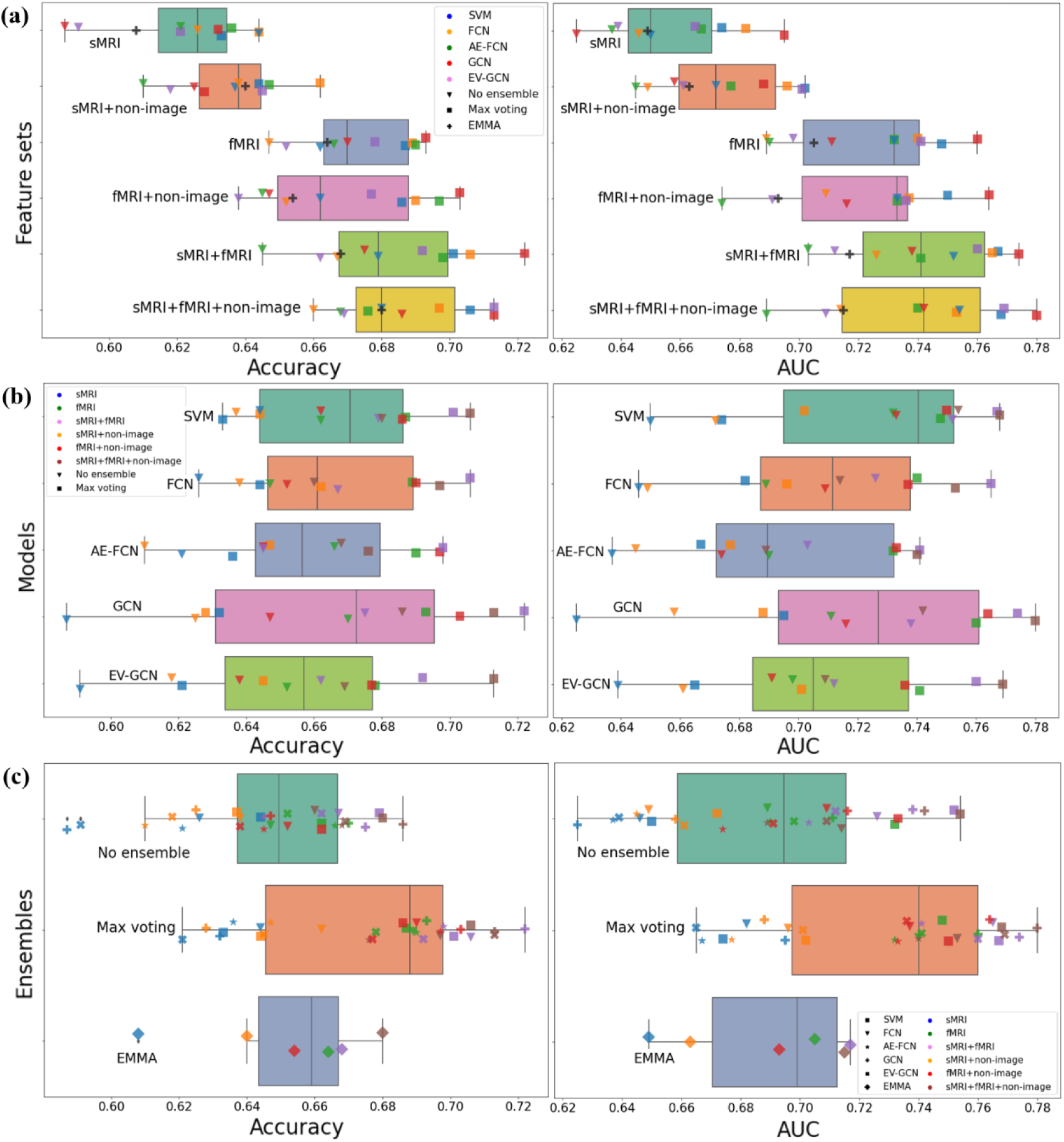
(a)The accuracies and AUC of different feature sets. (b) The accuracies and AUC of different machine learning models. (c) The accuracies and AUC of different ensemble situations.

**Table 5.** The experiment results of five machine learning models from (a) structural MRI (sMRI) features (b) sMRI + non-imaging features (c) functional MRI (fMRI) features (d) fMRI + non-imaging features (e) sMRI + fMRI features (f) sMRI + fMRI + non-imaging features. When ‘no ensemble’ is used, ‘validation’ is the best validation accuracy, ‘test’ is the corresponding test accuracy. AUC measures the area under the ROC curve of the predicted probability of the test set.

| (a) sMRI |  |  |  |  |  |  |  |
| --- | --- | --- | --- | --- | --- | --- | --- |
| CC200 | No ensemble |  |  | Max voting |  | EMMA |  |
|  | Validation | Test | AUC | Test | AUC | Test | AUC |
| SVM | 64.40% | 61.50% | 0.65 | 63.30% | 0.674 |  |  |
| FCN | 65.60% | 62.60% | 0.646 | **64.40%** | 0.682 |  |  |
| AE-FCN | 66.00% | 62.10% | 0.637 | 63.60% | 0.667 | 60.8% | 0.649 |
| GCN | 64.00% | 58.70% | 0.625 | 63.20% | **0.695** |  |  |
| EV-GCN | 65.70% | 59.10% | 0.639 | 62.10% | 0.665 |  |  |
| (b) sMRI + non-imaging |  |  |  |  |  |  |  |
| sMRI | No ensemble |  |  | Max voting |  | EMMA |  |
|  | Validation | Test | AUC | Test | AUC | Test | AUC |
| SVM | 66.10% | 63.70% | 0.672 | 64.40% | **0.702** |  |  |
| FCN | 66.10% | 63.80% | 0.649 | **66.20%** | 0.696 |  |  |
| AE-FCN | 67.40% | 61.00% | 0.645 | 64.70% | 0.677 | 64.0% | 0.663 |
| GCN | 64.60% | 62.50% | 0.658 | 62.80% | 0.688 |  |  |
| EV-GCN | 67.00% | 61.80% | 0.661 | 64.50% | 0.701 |  |  |

| (c) fMRI |  |  |  |  |  |  |  |
| --- | --- | --- | --- | --- | --- | --- | --- |
| sMRI | No ensemble |  |  | Max voting |  | EMMA |  |
|  | Validation | Test | AUC | Test | AUC | Test | AUC |
| SVM | 68.40% | 66.20% | 0.732 | 68.70% | 0.748 |  |  |
| FCN | 70.00% | 64.70% | 0.689 | 68.90% | 0.74 |  |  |
| AE-FCN | 70.80% | 66.60% | 0.69 | 69.00% | 0.732 | 66.4% | 0.705 |
| GCN | 70.80% | 67.00% | 0.711 | **69.30%** | **0.76** |  |  |
| EV-GCN | 68.70% | 65.20% | 0.698 | 67.80% | 0.741 |  |  |

| (d) fMRI + non-imaging |  |  |  |  |  |  |  |
| --- | --- | --- | --- | --- | --- | --- | --- |
| sMRI | No ensemble |  |  | Max voting |  | EMMA |  |
|  | Validation | Test | AUC | Test | AUC | Test | AUC |
| SVM | 68.60% | 66.20% | 0.733 | 68.60% | 0.75 |  |  |
| FCN | 70.60% | 65.20% | 0.709 | 69.00% | 0.737 |  |  |
| AE-FCN | 70.70% | 64.50% | 0.674 | 69.70% | 0.733 | 65.4% | 0.693 |
| GCN | 71.30% | 64.70% | 0.716 | **70.30%** | **0.764** |  |  |
| EV-GCN | 70.20% | 63.80% | 0.691 | 67.70% | 0.736 |  |  |

| (e) sMRI + fMRI |  |  |  |  |  |  |  |
| --- | --- | --- | --- | --- | --- | --- | --- |
| sMRI | No ensemble |  |  | Max voting |  | EMMA |  |
|  | Validation | Test | AUC | Test | AUC | Test | AUC |
| SVM | 69.90% | 67.90% | 0.752 | 70.10% | 0.767 |  |  |
| FCN | 71.30% | 66.70% | 0.726 | 70.60% | 0.765 |  |  |
| AE-FCN | 70.90% | 64.50% | 0.703 | 69.80% | 0.741 | 66.8% | 0.717 |
| GCN | 73.10% | 67.50% | 0.738 | **72.20%** | **0.774** |  |  |
| EV-GCN | 69.90% | 66.20% | 0.712 | 69.20% | 0.76 |  |  |

| (f) sMRI + fMRI + non-imaging |  |  |  |  |  |  |  |
| --- | --- | --- | --- | --- | --- | --- | --- |
| sMRI | No ensemble |  |  | Max voting |  | EMMA |  |
|  | Validation | Test | AUC | Test | AUC | Test | AUC |
| SVM | 70.10% | 68.00% | 0.754 | 70.60% | 0.768 |  |  |
| FCN | 72.41% | 66.00% | 0.714 | 69.70% | 0.753 |  |  |
| AE-FCN | 72.60% | 66.80% | 0.689 | 67.60% | 0.74 | 68.0% | 0.715 |
| GCN | 72.30% | 68.60% | 0.742 | **71.30%** | **0.78** |  |  |
| EV-GCN | 70.10% | 66.90% | 0.709 | **71.30%** | 0.769 |  |  |

#### 3.1.1 The overall performance of the models

Results presented in **Table 5** and **Fig. 8** show that the models achieved prediction accuracies on test set between 58% and 72%, and AUC between 0.64 and 0.78. The best performing model according to the test accuracy was the GCN with max-voting, trained on combined fMRI and sMRI features, achieving test accuracy 72.2% with the highest AUC value of 0.78 (see **Table 5e**).

#### 3.1.2 Predictive accuracy of different feature sets

To compare predictive ability of different feature sets, we present a boxplot of accuracies and AUC (**Fig. 8a**) for all models. We can observe a clear trend, where models trained on sMRI features have the lowest performance with an average accuracy of 62% and AUC of 0.65, while models trained on fMRI features achieved an average accuracy of 67% and AUC of 0.72. This was statistically significant (T-test, p =1.7e-6). Combining fMRI and sMRI features further improves the performance (average accuracy 68% and AUC 0.74), though this was not statistically significant (p=0.2). Adding non-imaging features did not result in a significant improvement in any of the feature sets. This suggests that functional connectivity plays a more prominent role in the prediction of autism diagnosis than structural features, as previously reported in the literature (Traut et al. 2022).

#### 3.1.3 Comparison of five machine learning models

The results presented in **Fig. 8b** demonstrate that all tested models performed similarly to classify Autism and TD on ABIDE dataset with P>0.05 when comparing each pair of models with T-test, indicating no significant improvement. Additionally, a Chi-squared test was conducted between the best performing model of each machine learning model, revealing no significant differences (P>0.05). This suggests that different inclusion criteria, data modalities and evaluation pipelines rather than different machine learning models cause the variation in accuracy in the published literature (Abraham et al. 2017; Heinsfeld et al. 2018; Huang and Chung 2020; Parisot et al. 2018; Rakić et al. 2020). High heterogeneity in term of collection site, image quality and biological heterogeneity of autism may limit the model performance.

Results in **Table 5** show that SVM, the only classical machine learning model tested as our baseline, achieved test accuracy 70.6% when used with sMRI+fMRI+non-imaging feature set and max voting. The performance is similar to best performing GCN, but the model is much faster and requires less computational resources than deep learning models. The performances of AE-FCN and EV-GCN are similar to other models, but are lower than the accuracies obtained by Huang and Chung (2020) and Rakić et al. (2020), which are 81% and 85% respectively. This difference in accuracy is most likely due to different inclusion criteria or evaluation strategies.

#### 3.1.4 Ensembles

Ensemble test accuracy using max voting (**Table 5** and **Fig. 8c)** enables the models to achieve better performance on the unseen dataset (test set), resulting in an average accuracy improvement of 3.8%, and this was significant over all models (p=0.0001). However, the results obtained with EMMA tended to outperform the average scores of the five models only slightly and, in some cases, even performed worse, with overall no significant difference with the individual models.

### 3.2 Characterisation of model stability using SmoothGrad saliency maps

The SNR value indicates whether a model assigns similar importance to the same features across various training sets. A high SNR value suggests that the model consistently relies on the same important features to classify participants into Autism or TD groups, regardless of the training set variability.

To compare the stability of GCN, FCN and AE-FCN models, we followed the diagram in **Fig. 7** to calculate the average SNR per participant of these 3 models and plotted the histogram in **Fig. 9**. The results illustrate that FCN model has the highest SNR values and therefore the most stable saliency maps across different training sets compared with GCN and AE-FCN. For this reason, we select the FCN model to extract important features from its average saliency map in section 3.3. Considering the best classification performance of FCN model is achieved on the sMRI + fMRI feature set, we will extract the most important features from this particular feature set.

**Fig. 9.**
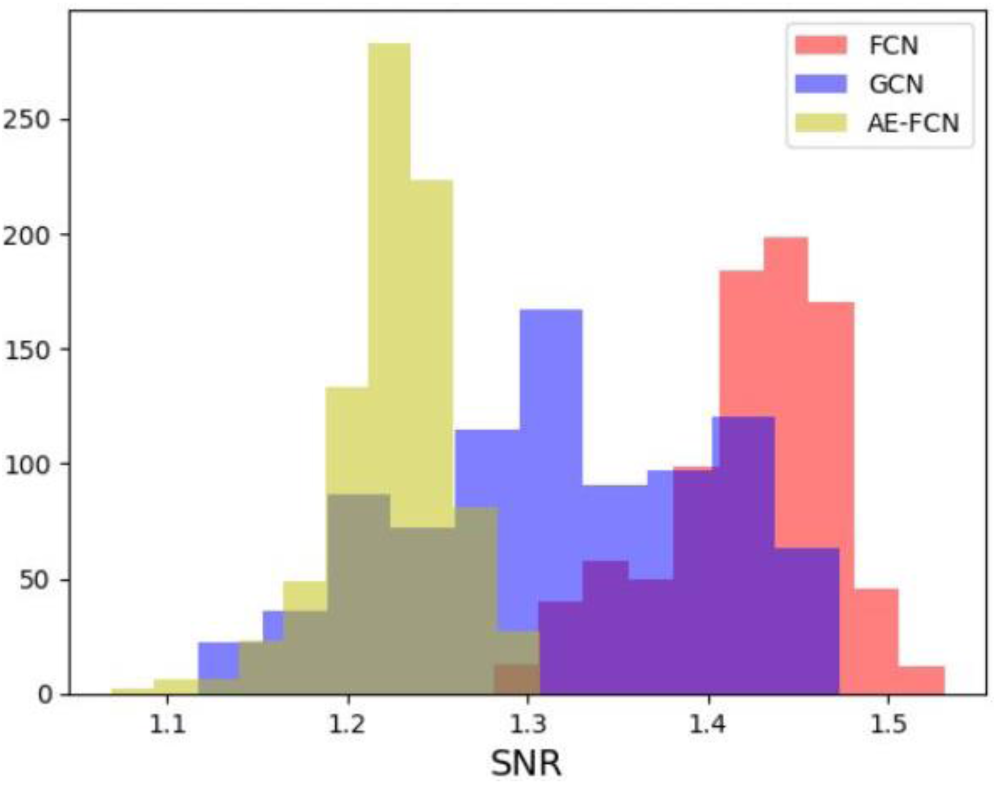
The SNR histograms of GCN, FCN and AE-FCN

### 3.3 Important features extracted from SmoothGrad saliency maps

We identified the significant fMRI and sMRI features from the FCN average saliency map based on saliency intensity. To better visualize and compare feature importance, we correlated the fMRI and sMRI saliency values with the regions defined in the CC200 atlas and the Desikan-Killiany atlas respectively, applied z-score normalization and further projected them onto a standardized brain template using the BrainNet Viewer software (Xia, Wang, and He 2013) (**Fig. 10**, **Fig. 11**).

**Fig. 10.**
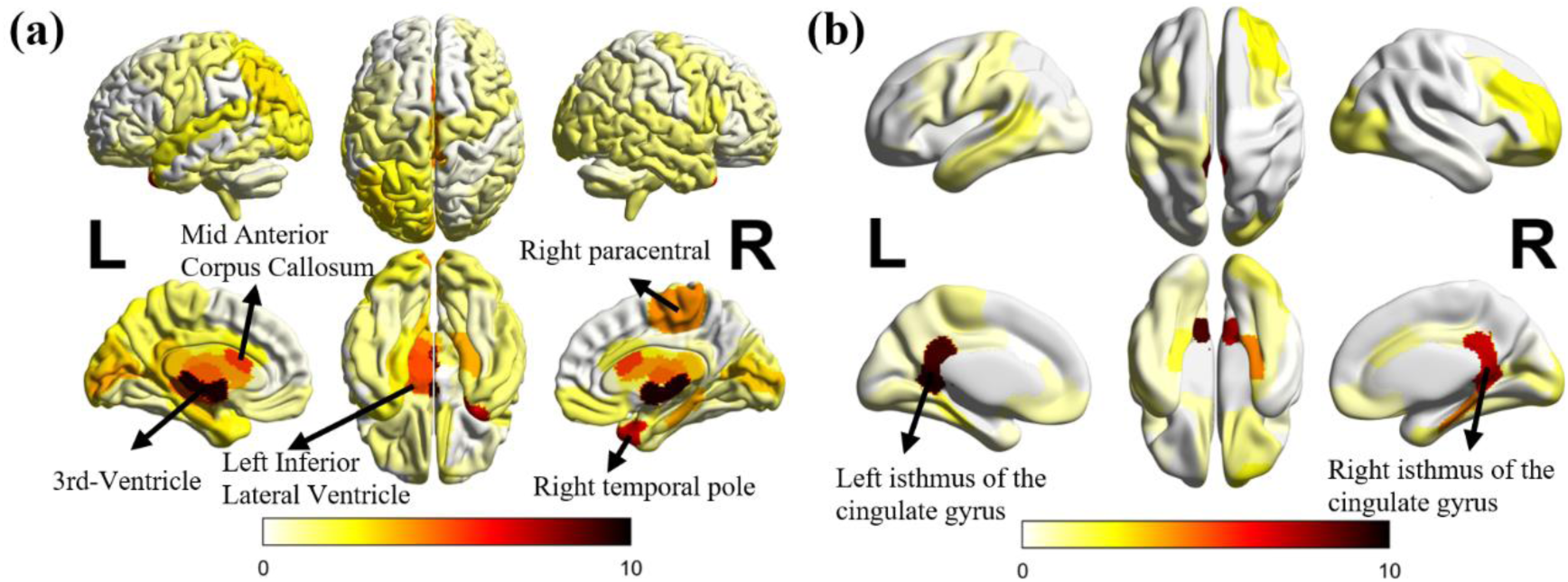
Important structural brain features that contribute to autism classification task. (a) Cortical + Subcortical areas. (b) White Matter areas.

**Fig. 11.**
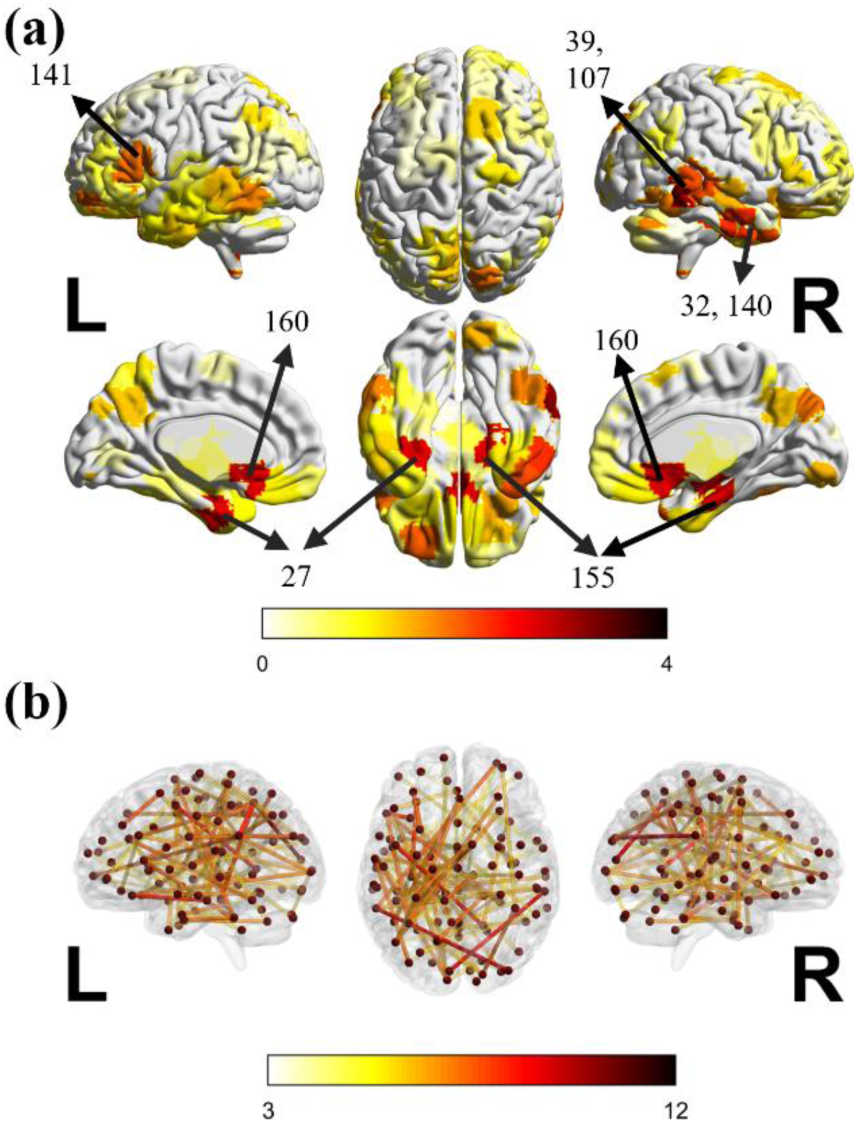
(a) The important brain regions obtained from fMRI features. The numbers indicate the corresponding brain regions in the CC200 atlas. The names of these brain regions are listed in the Supplementary **Table 1**. (b) Top 100 connections.

To assess the classification capability of the top selected features, we divided the entire population into four groups: true positive (TP, correctly classified as Autism), true negative (TN, correctly classified as TD), false positive (FP, incorrectly classified as Autism), and false negative (FN, incorrectly classified as TD). We then plotted box plots for each important feature’s normalized value across these four groups to compare the features’ value (**Fig. 12**).

**Fig. 12.**
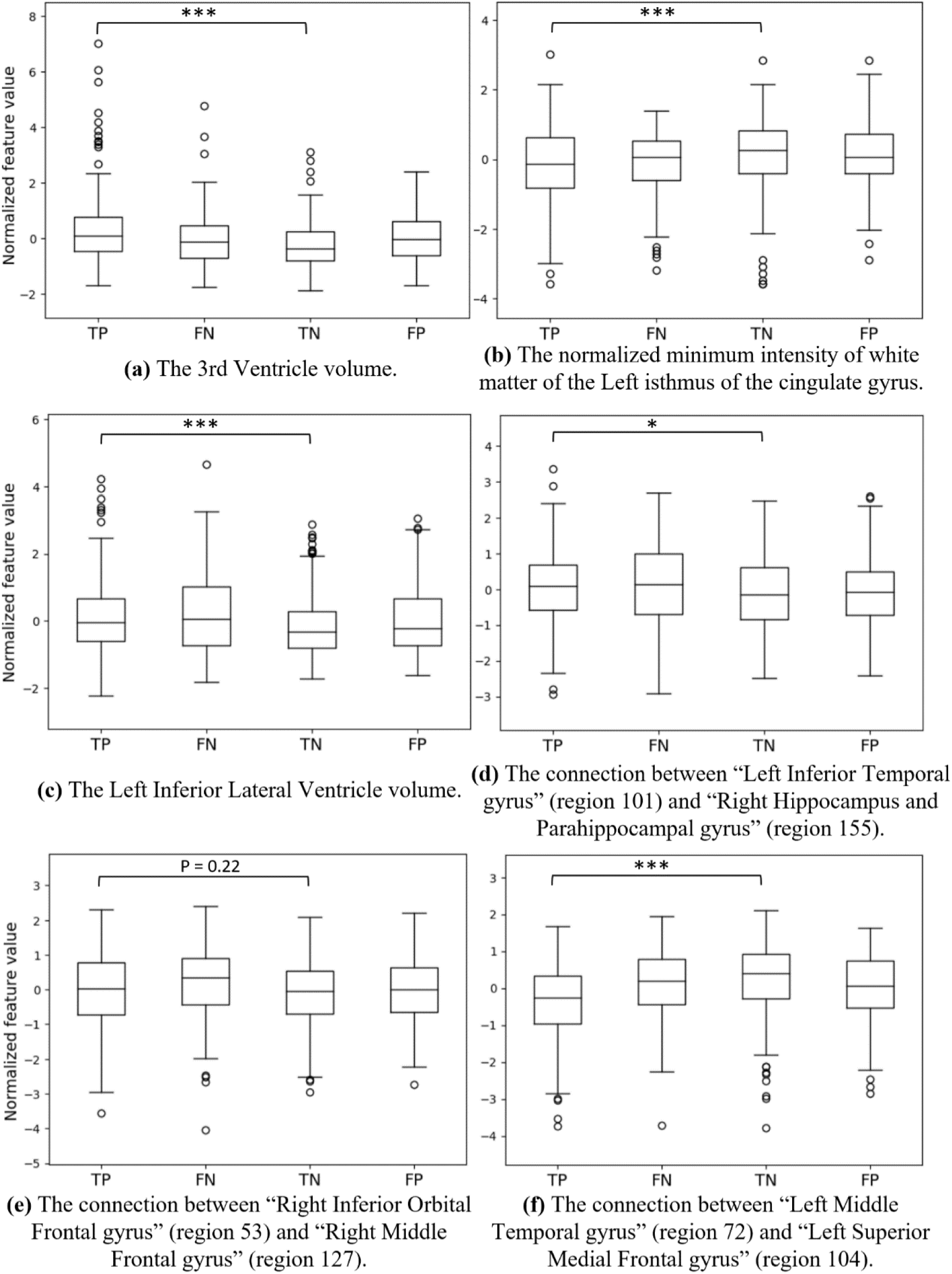
The box plots of the most important structural and functional features. P-value measures the difference between TP and TN groups. *: P<0.05, **: P<0.01, ***: P<0.001.

The top 100 most important structural volumetric features and brain connections are provided individually in the Supplementary **Table 1** and **Table 2**.

#### 3.3.1 Important sMRI features

We associated the saliency values of cortical, subcortical and white matter features with the Desikan-Killiany atlas and employed z-score normalization. Considering white matter areas are under cortical surface, we assigned the white matter saliency values to the corresponding cortical surface for better visualization and plotted Cortical and Subcortical areas (including ventricles)’ in **Fig. 10a**, and ‘White matter areas’ in **Fig. 10b**. A z-score below 0 indicates less importance to autism classification, whereas a score above 0 means greater importance to autism classification.

The regions with most relevant structural features identified by FCN were: 3^rd^ Ventricle, Left Inferior Lateral Ventricles, Right Paracentral lobule, and the white matter of Left and Right Isthmus Cingulate.

The box plots of three most important structural features are displayed in **Fig. 12a, b, c**, all showing significant differences between the TP and TN groups. These are the volume of the 3^rd^ Ventricle, the normalized minimum intensity of white matter of the Left isthmus of the cingulate gyrus, and the volume of the Left Inferior Lateral Ventricle. Notably, the boxplot of the feature with the highest z-score among all the structural features, the volume of the 3^rd^ Ventricle, shows that correctly classified autistic participants (TP) exhibit larger 3^rd^ Ventricle volume than the correctly classified TD participants (TN). The misclassified autistic and TD participants (FN and FP groups) respectively display smaller or larger volume size of the 3^rd^ Ventricle compared to TP and TN groups, probably contributing to the incorrect classification of these individuals.

#### 3.3.2 Important fMRI features

We assigned the saliency value of each functional connection to its corresponding pair of brain regions, summing up the values for each brain region (nodal saliency “strength”), and then applied z-score normalization. The most important brain regions used for classification of autistic individuals based on their functional connectivity, as well as specific functional brain connections used are visualized in **Fig. 11**.

The most significant brain regions selected by FCN model are located mostly in temporal cortex areas, including the Left Middle, Superior, and Inferior Temporal Cortex, as well as the Right Middle, Superior, and Inferior Temporal Cortex. Additionally, the Left Fusiform and Right Hippocampus and Parahippocampal regions were also highlighted as significant. This suggests that the connections associated with these brain regions may play a more substantial role in the Autism and TD classification compared to other connections.

The box plots of the top three connections are shown in **Fig. 12d, e, f**. Among them, two demonstrate a notable difference between TP and TN groups (p < 0.05), they are the connection between “Left Inferior Temporal gyrus” and “Right Hippocampus and Parahippocampal gyrus”, and the connection between “Left Middle Temporal gyrus” and “Left Superior Medial Frontal gyrus”.

## 4 Discussion

The purpose of this work was to evaluate the performance of five machine learning models classifying autistic individuals and TD participants under a standard setting, and extracting the features that contribute the most to the classification task with the most stable machine learning model. All the tested models showed similar classification performance, which illustrates that different inclusion criteria, data modalities and evaluation pipelines, rather than different machine learning models, cause variation in accuracy in published literature. Although the classical machine learning model SVM did not achieve the highest accuracy, it didn’t show significantly lower performance when compared with other more complex models. Our results suggests that, in this task, traditional machine learning models can produce similarly robust results to deep neural networks while being more computationally efficient.

The two evaluation pipelines utilized in this paper, 5-fold cross-validation and 5-fold nested cross-validation, were designed to provide robust and accurate results while also preventing the models from overfitting. Employing 5-fold cross-validation enables the evaluation of performance across all samples while ensuring that model training and tuning is independent of the test set (Cawley and Talbot 2010). The choice of 5 folds is a good compromise between robustness and the number of models that need to be trained. Additionally, it would be difficult to stratify the data into more folds due to the limited number of samples from some collection sites. We also note that performance calculated on the validation set (which we only show for all individual models with ‘no ensemble’) is always higher than on the test set. This shows the importance of reporting the performance on the test set to avoid unrealistically inflated performance measurements that would not repeat on unseen datasets. The application of max voting ensemble improved the performance of all five models compared to the individual models (‘no ensemble’). This shows that max voting can result in more robust models with stable performance on unseen datasets as previously suggested (Parisot et al. 2018; Rakić et al. 2020). While max voting consistently resulted in better performance, EMMA’s contribution was weaker, and in some cases even had a negative effect.

Our classification results show that feature sets used for training of the classifiers resulted in the most significant differences in model performance. We have observed that models trained with fMRI features (i.e., functional connectivity), exhibit better performance than the models trained with sMRI features. This suggests that functional connectivity plays a more prominent role in prediction of autism diagnosis than structural features, as previously reported in literature (Traut et al. 2022). When merging sMRI and fMRI features, the addition of sMRI information improved the performance only marginally. Moreover, adding non-imaging features did not yield a significant improvement in any of the feature sets.

Comprehensive experiments presented in this paper demonstrate that variations in experimental conditions, such as data modality and cross-validation technique, can lead to disparate outcomes, despite utilizing the same machine learning model. However, the differences in performance are moderate, with classification accuracy of around 70%. In fact, our results are consistent with most studies in the published literature that utilise the 871 samples of ABIDE fMRI dataset. This suggests that the limitations of this dataset, including inherent biological heterogeneity of autism, or site-dependent acquisition and processing characteristics, rather than the classification models, may be the primary cause of the limited classification performance. This highlights that fruitful directions of the future work will likely focus on better comprehension and disentanglement of the heterogeneity of autism and exploration of robust, effective stratification biomarkers.

FCN model combined with SmoothGrad yield the most stable interpretation results in comparison to other machine learning models. The lower SNR values observed for saliency features in GCN and AE-FCN models may be attributed to several factors. Firstly, the GCN model has the population graph as the second feature input, while FCN focuses on the functional and structural features only. Secondly, the AE-FCN model performs the classification and the denoising tasks at the same time, which could result in lower saliency SNR values.

Many studies have investigated atypical brain structural features in autism (Courchesne 2002; Katuwal et al. 2015; Moradi et al. 2017). In this study, we found the structural features in the ventricles contributed the most to the model decision-making. Some observations from our results are consistent with the published literature, for example, we observed an increased volume of the 3^rd^ Ventricle and Left and Right Lateral-Ventricle in autistic participants (p=1.3e-8), which aligns with findings from previous studies (Hardan et al. 2001; Palmen et al. 2005; Wolfe et al. 2015). The cortical features of the right paracentral lobule and the white matter near the isthmus cingulate gyrus also showed higher z-scores than other areas. These areas may play an important role in emotion regulation, emotion processing, and cognitive control in autism (Chien, Chen, and Gau 2021; Hau et al. 2019). Temporal cortex areas have been commonly associated with auditory perception, language and vision functions (Bonner and Price 2013; Hickok and Poeppel 2007; Zaehle et al. 2004). Alterations in functional connectivity within the temporal cortex may contribute to the auditory and language difficulties observed in individuals with autism (Rotschafer 2021; Xiao et al. 2023). It is worth noting that we didn’t identify a similar pattern of important brain regions for autism classification using functional and structural features, suggesting distinct patterns of regions with atypical structural morphology, and functional connectivity associated with autistic phenotypes.

One limitation of this study is that the samples used are solely from the ABIDE I dataset. In future, similar experiments may be performed on other datasets such as ABIDE II, and AIMS-2-TRIALS, (Loth et al. 2017). The investigation of longitudinal autism databases may enable the identification and development of early stratification biomarkers for autism, potentially leading to more effective personalised supportive strategies for those individuals that request them. Another limitation of our study is that we mainly utilized functional connectivity and sMRI volumetric features. Other types of features, such as topological properties of functional connectivity matrices (Kazeminejad and Sotero 2019; Plitt, Barnes, and Martin 2015), extra-axial cerebrospinal fluid (Shen et al. 2017) and genomic copy number variations (Sherman et al. 2021; Velinov 2019) also hold significant potential for autism classification and stratification.

## 5 Conclusion

In this paper, five machine learning models from the existing literature were trained to classify individuals with autism using ABIDE I dataset. The results showed that all tested models had similar performance, indicating that variations in accuracy in published literature may be attributed to different inclusion criteria, data modalities and evaluation pipelines rather than the models themselves. The highest classification performance was obtained by combining fMRI and sMRI features, to train the GCN model with max voting, resulting in a classification accuracy of 72.2% and AUC of 0.78. Ensemble using max voting method was found to consistently improve the performance of the models. We conclude that the performance of the classifiers is likely limited by other factors than the model architecture, such as high heterogeneity in terms of age, collection site and image quality, as well as biological heterogeneity of autism in general. Furthermore, SmoothGrad method was applied to FCN, which exhibited the highest SNR value and was selected as the most stable model. The results suggested that structural features from the ventricles and functional features from the temporal cortex made the most significant contributions to the algorithms identifying autistic participants.

## Data Availability

All data used in the study are available online at the ABIDE dataset repository http://fcon_1000.projects.nitrc.org/indi/abide/

http://preprocessed-connectomes-project.org/abide/

## Acknowledgements

This work was supported by King’s-China Scholarship Council (K-CSC) [grant number 202008060109]. The authors acknowledge support in part from the Wellcome Engineering and Physical Sciences Research Council (EPSRC) Centre for Medical Engineering at Kings College London [WT 203148/Z/16/Z], and the NIHR Maudsley Biomedical Research Centre at South London and Maudsley NHS Foundation Trust and King’s College London. The views expressed are those of the author(s) and not necessarily those of the NIHR or the Department of Health and Social Care.

## Conflict of Interest Statement

The authors declare that they have no known competing financial interests or personal relationships that could have appeared to influence the work reported in this paper.

## Notes

### Competing Interest Statement

The authors have declared no competing interest.

### Author Declarations

ABIDE database: https://fcon_1000.projects.nitrc.org/indi/abide/ Data acquire: http://preprocessed-connectomes-project.org/abide/

