## Supplementary Table 1 Supplementary Table 2 for "Reproducible comparison and interpretation of machine learning classifiers to predict autism on the ABIDE multimodal dataset"

Supplementary Table 1. The top 100 most important brain connections

| The top 100 connections | Brain regions in CC200 atlas | The corresponding brain regions in AAL atlas |
| --- | --- | --- |
| 1 | 101 | Temporal_Inf_L: 0.58, Temporal_Mid_L: 0.38 |
|  | 155 | Hippocampus_R: 0.39, ParaHippocampal_R: 0.37, None: 0.11 |
| 2 | 53 | Frontal_Inf_Orb_R: 0.88 |
|  | 127 | Frontal_Mid_R: 0.70, Frontal_Sup_R: 0.30 |
| 3 | 72 | Temporal_Mid_L: 0.88 |
|  | 104 | Frontal_Sup_Medial_L: 0.70, Frontal_Sup_L: 0.29 |
| 4 | 32 | Temporal_Pole_Mid_R: 0.59, Temporal_Inf_R: 0.35 |
|  | 155 | Hippocampus_R: 0.39, ParaHippocampal_R: 0.37, None: 0.11 |
| 5 | 56 | Parietal_Inf_L: 0.85 |
|  | 144 | Frontal_Inf_Tri_R: 0.91 |
| 6 | 19 | Calcarine_L: 0.37, Cuneus_L: 0.36, Precuneus_L: 0.16 |
|  | 140 | Temporal_Mid_R: 0.65, Temporal_Inf_R: 0.33 |
| 7 | 155 | Hippocampus_R: 0.39, ParaHippocampal_R: 0.37, None: 0.11 |
|  | 198 | Fusiform_R: 0.41, ParaHippocampal_R: 0.25, Temporal_Pole_Mid_R: 0.15, Temporal_Inf_R: 0.13 |
| 8 | 17 | Frontal_Inf_Oper_R: 0.44, Precentral_R: 0.31, Frontal_Mid_R: 0.13, Frontal_Inf_Tri_R: 0.13 |
|  | 20 | Insula_L: 0.76, Frontal_Inf_Tri_L: 0.12 |
| 9 | 40 | Cingulum_Ant_R: 0.44, Cingulum_Ant_L: 0.40 |
|  | 140 | Temporal_Mid_R: 0.65, Temporal_Inf_R: 0.33 |
| 10 | 102 | Occipital_Mid_R: 1.00 |
|  | 160 | Olfactory_L: 0.27, Olfactory_R: 0.22 |
| 11 | 26 | Occipital_Inf_R: 0.55, Temporal_Inf_R: 0.16, Cerebelum_Crus1_R: 0.11 |
|  | 102 | Occipital_Mid_R: 1.00 |
| 12 | 94 | Caudate_R: 0.84, Thalamus_R: 0.14 |
|  | 124 | Frontal_Mid_Orb_R: 0.61, Frontal_Sup_Orb_R: 0.28 |
| 13 | 11 | Temporal_Mid_L: 0.96 |
|  | 32 | Temporal_Pole_Mid_R: 0.59, Temporal_Inf_R: 0.35 |
| 14 | 92 | Hippocampus_L: 0.41, Amygdala_L: 0.29, ParaHippocampal_L: 0.15 |
|  | 141 | Frontal_Inf_Tri_L: 0.96 |
| 15 | 145 | Hippocampus_L: 0.38, Fusiform_L: 0.31, ParaHippocampal_L: 0.16, Temporal_Inf_L: 0.15 |
|  | 199 | ParaHippocampal_L: 0.38, Cerebelum_4_5_L: 0.34, Cerebelum_3_L: 0.18 |
| 16 | 140 | Temporal_Mid_R: 0.65, Temporal_Inf_R: 0.33 |
|  | 155 | Hippocampus_R: 0.39, ParaHippocampal_R: 0.37, None: 0.11 |
| 17 | 39 | Temporal_Mid_R: 0.67, Temporal_Inf_R: 0.33 |
|  | 155 | Hippocampus_R: 0.39, ParaHippocampal_R: 0.37, None: 0.11 |
| 18 | 47 | Caudate_L: 0.45, Putamen_L: 0.39 |
|  | 135 | Caudate_R: 0.85, Putamen_R: 0.11 |
| 19 | 87 | Fusiform_R: 0.32, ParaHippocampal_R: 0.23, Temporal_Inf_R: 0.23, Hippocampus_R: 0.22 |
|  | 96 | Postcentral_L: 0.65, Precentral_L: 0.21, Parietal_Sup_L: 0.12 |
| 20 | 70 | Calcarine_L: 0.51, Lingual_L: 0.49 |
|  | 81 | Cuneus_R: 0.51, Precuneus_R: 0.31, Occipital_Sup_R: 0.14 |
| 21 | 39 | Temporal_Mid_R: 0.67, Temporal_Inf_R: 0.33 |
|  | 62 | Fusiform_R: 0.54, Hippocampus_R: 0.21, ParaHippocampal_R: 0.19 |
| 22 | 100 | Temporal_Inf_R: 0.66, Fusiform_R: 0.26 |
|  | 189 | Fusiform_L: 0.51, Cerebelum_6_L: 0.29, Lingual_L: 0.17 |
| 23 | 95 | Frontal_Mid_L: 0.72, Frontal_Sup_L: 0.28 |
|  | 147 | Precuneus_L: 0.54, Cuneus_L: 0.33 |
| 24 | 56 | Parietal_Inf_L: 0.85 |
|  | 107 | Temporal_Sup_R: 0.50, Temporal_Mid_R: 0.49 |
| 25 | 107 | Temporal_Sup_R: 0.50, Temporal_Mid_R: 0.49 |
|  | 153 | Temporal_Sup_R: 0.64, Temporal_Mid_R: 0.33 |
| 26 | 81 | Cuneus_R: 0.51, Precuneus_R: 0.31, Occipital_Sup_R: 0.14 |
|  | 105 | Lingual_R: 0.53, Calcarine_R: 0.31 |
| 27 | 99 | Temporal_Mid_L: 0.60, Temporal_Inf_L: 0.40 |
|  | 151 | Frontal_Inf_Tri_L: 0.73, Frontal_Mid_L: 0.20 |
| 28 | 26 | Occipital_Inf_R: 0.55, Temporal_Inf_R: 0.16, Cerebelum_Crus1_R: 0.11 |
|  | 175 | Occipital_Inf_R: 0.52, Fusiform_R: 0.15, Lingual_R: 0.14, Cerebelum_Crus1_R: 0.14 |
| 29 | 63 | Temporal_Inf_L: 0.45, Fusiform_L: 0.36, Cerebelum_Crus1_L: 0.13 |
|  | 197 | Precuneus_L: 0.92 |
| 30 | 48 | Hippocampus_R: 0.28, Lingual_R: 0.21, Thalamus_R: 0.20, ParaHippocampal_R: 0.15 |
|  | 199 | ParaHippocampal_L: 0.38, Cerebelum_4_5_L: 0.34, Cerebelum_3_L: 0.18 |
| 31 | 128 | SupraMarginal_R: 0.82, Temporal_Sup_R: 0.17 |
|  | 188 | Parietal_Sup_L: 0.61, Precuneus_L: 0.37 |
| 32 | 40 | Cingulum_Ant_R: 0.44, Cingulum_Ant_L: 0.40 |
|  | 72 | Temporal_Mid_L: 0.88 |
| 33 | 104 | Frontal_Sup_Medial_L: 0.70, Frontal_Sup_L: 0.29 |
|  | 109 | Frontal_Med_Orb_L: 0.37, Frontal_Med_Orb_R: 0.35 |
| 34 | 107 | Temporal_Sup_R: 0.50, Temporal_Mid_R: 0.49 |
|  | 150 | Occipital_Mid_L: 0.54, Temporal_Mid_L: 0.46 |
| 35 | 20 | Insula_L: 0.76, Frontal_Inf_Tri_L: 0.12 |
|  | 144 | Frontal_Inf_Tri_R: 0.91 |
| 36 | 32 | Temporal_Pole_Mid_R: 0.59, Temporal_Inf_R: 0.35 |
|  | 179 | Lingual_L: 0.48, Cerebelum_4_5_L: 0.21, Calcarine_L: 0.14, Precuneus_L: 0.12 |
| 37 | 87 | Fusiform_R: 0.32, ParaHippocampal_R: 0.23, Temporal_Inf_R: 0.23, Hippocampus_R: 0.22 |
|  | 122 | Fusiform_L: 0.47, ParaHippocampal_L: 0.23, Hippocampus_L: 0.21 |
| 38 | 140 | Temporal_Mid_R: 0.65, Temporal_Inf_R: 0.33 |
|  | 174 | Precuneus_L: 0.46, Precuneus_R: 0.36 |
| 39 | 160 | Olfactory_L: 0.27, Olfactory_R: 0.22 |
|  | 170 | Occipital_Mid_R: 0.70, Occipital_Sup_R: 0.30 |
| 40 | 74 | Frontal_Inf_Orb_L: 0.90 |
|  | 92 | Hippocampus_L: 0.41, Amygdala_L: 0.29, ParaHippocampal_L: 0.15 |
| 41 | 81 | Cuneus_R: 0.51, Precuneus_R: 0.31, Occipital_Sup_R: 0.14 |
|  | 95 | Frontal_Mid_L: 0.72, Frontal_Sup_L: 0.28 |
| 42 | 47 | Caudate_L: 0.45, Putamen_L: 0.39 |
|  | 94 | Caudate_R: 0.84, Thalamus_R: 0.14 |
| 43 | 42 | Frontal_Mid_Orb_L: 0.78, Frontal_Inf_Orb_L: 0.14 |
|  | 78 | Temporal_Pole_Mid_L: 0.54, Temporal_Pole_Sup_L: 0.40 |
| 44 | 141 | Frontal_Inf_Tri_L: 0.96 |
|  | 155 | Hippocampus_R: 0.39, ParaHippocampal_R: 0.37, None: 0.11 |
| 45 | 39 | Temporal_Mid_R: 0.67, Temporal_Inf_R: 0.33 |
|  | 86 | Cerebelum_9_R: 0.48, None: 0.29, Vermis_10: 0.14 |
| 46 | 59 | Insula_R: 0.61, Putamen_R: 0.20, Frontal_Inf_Tri_R: 0.11 |
|  | 144 | Frontal_Inf_Tri_R: 0.91 |
| 47 | 75 | Frontal_Mid_R: 0.58, Frontal_Sup_R: 0.42 |
|  | 196 | Temporal_Pole_Sup_L: 0.62, Frontal_Inf_Orb_L: 0.22, Insula_L: 0.11 |
| 48 | 63 | Temporal_Inf_L: 0.45, Fusiform_L: 0.36, Cerebelum_Crus1_L: 0.13 |
|  | 136 | Parietal_Sup_L: 0.50, Precuneus_L: 0.50 |
| 49 | 50 | Precentral_L: 0.64, Frontal_Sup_L: 0.27 |
|  | 150 | Occipital_Mid_L: 0.54, Temporal_Mid_L: 0.46 |
| 50 | 141 | Frontal_Inf_Tri_L: 0.96 |
|  | 145 | Hippocampus_L: 0.38, Fusiform_L: 0.31, ParaHippocampal_L: 0.16, Temporal_Inf_L: 0.15 |
| 51 | 49 | Temporal_Mid_R: 0.64, Temporal_Inf_R: 0.34 |
|  | 62 | Fusiform_R: 0.54, Hippocampus_R: 0.21, ParaHippocampal_R: 0.19 |
| 52 | 81 | Cuneus_R: 0.51, Precuneus_R: 0.31, Occipital_Sup_R: 0.14 |
|  | 89 | Calcarine_R: 0.43, Lingual_R: 0.35 |
| 53 | 100 | Temporal_Inf_R: 0.66, Fusiform_R: 0.26 |
|  | 102 | Occipital_Mid_R: 1.00 |
| 54 | 26 | Occipital_Inf_R: 0.55, Temporal_Inf_R: 0.16, Cerebelum_Crus1_R: 0.11 |
|  | 160 | Olfactory_L: 0.27, Olfactory_R: 0.22 |
| 55 | 4 | Temporal_Sup_L: 0.33, Putamen_L: 0.28, Insula_L: 0.25 |
|  | 198 | Fusiform_R: 0.41, ParaHippocampal_R: 0.25, Temporal_Pole_Mid_R: 0.15, Temporal_Inf_R: 0.13 |
| 56 | 6 | Cingulum_Mid_R: 0.60, Precuneus_R: 0.21 |
|  | 147 | Precuneus_L: 0.54, Cuneus_L: 0.33 |
| 57 | 63 | Temporal_Inf_L: 0.45, Fusiform_L: 0.36, Cerebelum_Crus1_L: 0.13 |
|  | 168 | Frontal_Mid_R: 0.96 |
| 58 | 58 | Precuneus_L: 0.31, Precuneus_R: 0.28, Vermis_4_5: 0.14 |
|  | 109 | Frontal_Med_Orb_L: 0.37, Frontal_Med_Orb_R: 0.35 |
| 59 | 76 | Cingulum_Mid_L: 0.86 |
|  | 107 | Temporal_Sup_R: 0.50, Temporal_Mid_R: 0.49 |
| 60 | 119 | Rolandic_Oper_R: 0.42, Frontal_Inf_Oper_R: 0.23, Temporal_Sup_R: 0.19 |
|  | 188 | Parietal_Sup_L: 0.61, Precuneus_L: 0.37 |
| 61 | 14 | Angular_R: 0.46, Temporal_Mid_R: 0.36 |
|  | 81 | Cuneus_R: 0.51, Precuneus_R: 0.31, Occipital_Sup_R: 0.14 |
| 62 | 11 | Temporal_Mid_L: 0.96 |
|  | 78 | Temporal_Pole_Mid_L: 0.54, Temporal_Pole_Sup_L: 0.40 |
| 63 | 65 | Postcentral_R: 0.33, Paracentral_Lobule_R: 0.26, Precuneus_R: 0.22, Parietal_Sup_R: 0.19 |
|  | 96 | Postcentral_L: 0.65, Precentral_L: 0.21, Parietal_Sup_L: 0.12 |
| 64 | 57 | Frontal_Inf_Orb_L: 0.72, Frontal_Mid_Orb_L: 0.25 |
|  | 127 | Frontal_Mid_R: 0.70, Frontal_Sup_R: 0.30 |
| 65 | 25 | Frontal_Mid_R: 0.69, Frontal_Inf_Tri_R: 0.31 |
|  | 183 | Frontal_Mid_L: 0.43, Frontal_Sup_L: 0.42 |
| 66 | 39 | Temporal_Mid_R: 0.67, Temporal_Inf_R: 0.33 |
|  | 58 | Precuneus_L: 0.31, Precuneus_R: 0.28, Vermis_4_5: 0.14 |
| 67 | 71 | Frontal_Inf_Orb_R: 0.55, Insula_R: 0.30 |
|  | 112 | Insula_L: 0.34, Frontal_Inf_Orb_L: 0.29, Temporal_Pole_Sup_L: 0.26 |
| 68 | 153 | Temporal_Sup_R: 0.64, Temporal_Mid_R: 0.33 |
|  | 194 | None: 0.71, Vermis_3: 0.14 |
| 69 | 38 | Frontal_Mid_R: 0.77, Frontal_Inf_Tri_R: 0.14 |
|  | 113 | Frontal_Mid_Orb_R: 0.42, Frontal_Mid_R: 0.28, Frontal_Inf_Orb_R: 0.25 |
| 70 | 160 | Olfactory_L: 0.27, Olfactory_R: 0.22 |
|  | 175 | Occipital_Inf_R: 0.52, Fusiform_R: 0.15, Lingual_R: 0.14, Cerebelum_Crus1_R: 0.14 |
| 71 | 32 | Temporal_Pole_Mid_R: 0.59, Temporal_Inf_R: 0.35 |
|  | 97 | Occipital_Mid_L: 0.87, Occipital_Sup_L: 0.11 |
| 72 | 144 | Frontal_Inf_Tri_R: 0.91 |
|  | 153 | Temporal_Sup_R: 0.64, Temporal_Mid_R: 0.33 |
| 73 | 27 | Fusiform_L: 0.45, Temporal_Inf_L: 0.21, ParaHippocampal_L: 0.13 |
|  | 57 | Frontal_Inf_Orb_L: 0.72, Frontal_Mid_Orb_L: 0.25 |
| 74 | 19 | Calcarine_L: 0.37, Cuneus_L: 0.36, Precuneus_L: 0.16 |
|  | 147 | Precuneus_L: 0.54, Cuneus_L: 0.33 |
| 75 | 51 | Frontal_Med_Orb_L: 0.27, Rectus_L: 0.23, Frontal_Med_Orb_R: 0.21, Rectus_R: 0.18 |
|  | 103 | Cerebelum_6_L: 0.46, Cerebelum_4_5_L: 0.32 |
| 76 | 10 | Cerebelum_Crus1_R: 0.77, Cerebelum_6_R: 0.11 |
|  | 117 | Temporal_Mid_L: 0.82, Temporal_Sup_L: 0.18 |
| 77 | 23 | Frontal_Inf_Tri_L: 0.57, Frontal_Mid_L: 0.42 |
|  | 42 | Frontal_Mid_Orb_L: 0.78, Frontal_Inf_Orb_L: 0.14 |
| 78 | 39 | Temporal_Mid_R: 0.67, Temporal_Inf_R: 0.33 |
|  | 99 | Temporal_Mid_L: 0.60, Temporal_Inf_L: 0.40 |
| 79 | 23 | Frontal_Inf_Tri_L: 0.57, Frontal_Mid_L: 0.42 |
|  | 91 | Frontal_Sup_Medial_L: 0.56, Frontal_Sup_Medial_R: 0.34 |
| 80 | 39 | Temporal_Mid_R: 0.67, Temporal_Inf_R: 0.33 |
|  | 97 | Occipital_Mid_L: 0.87, Occipital_Sup_L: 0.11 |
| 81 | 70 | Calcarine_L: 0.51, Lingual_L: 0.49 |
|  | 164 | Frontal_Inf_Tri_R: 0.62, Frontal_Inf_Oper_R: 0.36 |
| 82 | 42 | Frontal_Mid_Orb_L: 0.78, Frontal_Inf_Orb_L: 0.14 |
|  | 43 | Temporal_Inf_L: 0.50, Temporal_Pole_Mid_L: 0.26, None: 0.16 |
| 83 | 18 | Thalamus_R: 0.99 |
|  | 107 | Temporal_Sup_R: 0.50, Temporal_Mid_R: 0.49 |
| 84 | 20 | Insula_L: 0.76, Frontal_Inf_Tri_L: 0.12 |
|  | 39 | Temporal_Mid_R: 0.67, Temporal_Inf_R: 0.33 |
| 85 | 155 | Hippocampus_R: 0.39, ParaHippocampal_R: 0.37, None: 0.11 |
|  | 193 | Frontal_Sup_R: 0.50, Frontal_Sup_Medial_R: 0.42 |
| 86 | 133 | Frontal_Sup_Medial_L: 0.62, Frontal_Sup_L: 0.36 |
|  | 155 | Hippocampus_R: 0.39, ParaHippocampal_R: 0.37, None: 0.11 |
| 87 | 54 | Cuneus_L: 0.62, Occipital_Sup_L: 0.20 |
|  | 129 | Temporal_Sup_L: 0.34, Temporal_Mid_L: 0.26, Temporal_Pole_Sup_L: 0.17 |
| 88 | 87 | Fusiform_R: 0.32, ParaHippocampal_R: 0.23, Temporal_Inf_R: 0.23, Hippocampus_R: 0.22 |
|  | 179 | Lingual_L: 0.48, Cerebelum_4_5_L: 0.21, Calcarine_L: 0.14, Precuneus_L: 0.12 |
| 89 | 129 | Temporal_Sup_L: 0.34, Temporal_Mid_L: 0.26, Temporal_Pole_Sup_L: 0.17 |
|  | 140 | Temporal_Mid_R: 0.65, Temporal_Inf_R: 0.33 |
| 90 | 118 | Cerebelum_6_R: 0.70, Cerebelum_4_5_R: 0.18 |
|  | 166 | Angular_R: 0.85, Occipital_Mid_R: 0.13 |
| 91 | 75 | Frontal_Mid_R: 0.58, Frontal_Sup_R: 0.42 |
|  | 106 | Frontal_Mid_R: 0.72, Frontal_Sup_R: 0.28 |
| 92 | 20 | Insula_L: 0.76, Frontal_Inf_Tri_L: 0.12 |
|  | 115 | Precentral_R: 0.56, Frontal_Mid_R: 0.44 |
| 93 | 6 | Cingulum_Mid_R: 0.60, Precuneus_R: 0.21 |
|  | 81 | Cuneus_R: 0.51, Precuneus_R: 0.31, Occipital_Sup_R: 0.14 |
| 94 | 11 | Temporal_Mid_L: 0.96 |
|  | 51 | Frontal_Med_Orb_L: 0.27, Rectus_L: 0.23, Frontal_Med_Orb_R: 0.21, Rectus_R: 0.18 |
| 95 | 27 | Fusiform_L: 0.45, Temporal_Inf_L: 0.21, ParaHippocampal_L: 0.13 |
|  | 114 | Occipital_Mid_L: 0.32, Occipital_Sup_L: 0.27, Parietal_Sup_L: 0.26, Parietal_Inf_L: 0.15 |
| 96 | 114 | Occipital_Mid_L: 0.32, Occipital_Sup_L: 0.27, Parietal_Sup_L: 0.26, Parietal_Inf_L: 0.15 |
|  | 160 | Olfactory_L: 0.27, Olfactory_R: 0.22 |
| 97 | 32 | Temporal_Pole_Mid_R: 0.59, Temporal_Inf_R: 0.35 |
|  | 178 | Putamen_R: 0.69, Pallidum_R: 0.21 |
| 98 | 140 | Temporal_Mid_R: 0.65, Temporal_Inf_R: 0.33 |
|  | 178 | Putamen_R: 0.69, Pallidum_R: 0.21 |
| 99 | 57 | Frontal_Inf_Orb_L: 0.72, Frontal_Mid_Orb_L: 0.25 |
|  | 122 | Fusiform_L: 0.47, ParaHippocampal_L: 0.23, Hippocampus_L: 0.21 |
| 100 | 32 | Temporal_Pole_Mid_R: 0.59, Temporal_Inf_R: 0.35 |
|  | 106 | Frontal_Mid_R: 0.72, Frontal_Sup_R: 0.28 |

Supplementary Table 2. The top 100 most important structural volumetric features

| The top 100 structural features | Desikan-Killiany atlas regions | feature names |
| --- | --- | --- |
| 1 | 3rd-Ventricle | Volume |
| 2 | wm-lh-isthmuscingulate | Intensity normMin |
| 3 | Left-Inf-Lat-Vent | Volume |
| 4 | 3rd-Ventricle | Number of Voxels |
| 5 | wm-rh-isthmuscingulate | Intensity normRange |
| 6 | ctx-rh-temporalpole | CurvInd |
| 7 | wm-rh-isthmuscingulate | Intensity normMin |
| 8 | CC_Mid_Anterior | Intensity normMin |
| 9 | wm-lh-isthmuscingulate | Intensity normRange |
| 10 | wm-lh-isthmuscingulate | Itensity normStdDev |
| 11 | Left-VentralDC | Volume |
| 12 | Left-Thalamus | Intensity normMax |
| 13 | Left-Inf-Lat-Vent | Number of Voxels |
| 14 | 4th-Ventricle | Itensity normStdDev |
| 15 | wm-rh-parahippocampal | Volume |
| 16 | CSF | Volume |
| 17 | ctx-rh-paracentral | ThickAvg |
| 18 | Left-VentralDC | Number of Voxels |
| 19 | Left-choroid-plexus | Intensity normMean |
| 20 | non-WM-hypointensities | Itensity normStdDev |
| 21 | Left-Lateral-Ventricle | Itensity normStdDev |
| 22 | ctx-lh-pericalcarine | MeanCurv |
| 23 | CC_Mid_Anterior | Intensity normRange |
| 24 | ctx-rh-parahippocampal | NumVert |
| 25 | Left-Caudate | Intensity normMax |
| 26 | CSF | Number of Voxels |
| 27 | CC_Posterior | Intensity normMin |
| 28 | ctx-rh-pericalcarine | MeanCurv |
| 29 | Left-Pallidum | Intensity normMax |
| 30 | ctx-lh-isthmuscingulate | MeanCurv |
| 31 | ctx-lh-cuneus | GausCurv |
| 32 | wm-rh-parahippocampal | Number of Voxels |
| 33 | Left-Hippocampus | Itensity normStdDev |
| 34 | ctx-lh-inferiorparietal | FoldInd |
| 35 | ctx-lh-entorhinal | FoldInd |
| 36 | ctx-lh-pericalcarine | GausCurv |
| 37 | ctx-rh-parahippocampal | GrayVol |
| 38 | CC_Central | Volume |
| 39 | ctx-lh-pericalcarine | CurvInd |
| 40 | ctx-rh-pericalcarine | ThickStd |
| 41 | wm-rh-rostralmiddlefrontal | Intensity normMin |
| 42 | ctx-lh-superiortemporal | ThickAvg |
| 43 | ctx-lh-caudalanteriorcingulate | FoldInd |
| 44 | ctx-lh-superiorparietal | CurvInd |
| 45 | ctx-lh-parahippocampal | NumVert |
| 46 | ctx-rh-temporalpole | FoldInd |
| 47 | ctx-lh-entorhinal | ThickStd |
| 48 | ctx-lh-temporalpole | ThickStd |
| 49 | ctx-lh-lateraloccipital | ThickAvg |
| 50 | Left-Lateral-Ventricle | Volume |
| 51 | ctx-lh-parahippocampal | CurvInd |
| 52 | CC_Mid_Posterior | Intensity normMin |
| 53 | wm-lh-isthmuscingulate | Number of Voxels |
| 54 | wm-lh-parahippocampal | Volume |
| 55 | Left-Lateral-Ventricle | Number of Voxels |
| 56 | ctx-lh-entorhinal | CurvInd |
| 57 | ctx-rh-caudalanteriorcingulate | GausCurv |
| 58 | WM-hypointensities | Intensity normMean |
| 59 | Right-Inf-Lat-Vent | Itensity normStdDev |
| 60 | ctx-lh-posteriorcingulate | MeanCurv |
| 61 | ctx-lh-parahippocampal | SurfArea |
| 62 | wm-lh-bankssts | Volume |
| 63 | Left-Hippocampus | Intensity normMax |
| 64 | wm-lh-bankssts | Itensity normStdDev |
| 65 | Right-UnsegmentedWhiteMatter | Itensity normStdDev |
| 66 | ctx-rh-parahippocampal | SurfArea |
| 67 | wm-rh-rostralmiddlefrontal | Intensity normRange |
| 68 | CC_Central | Number of Voxels |
| 69 | ctx-rh-parsopercularis | MeanCurv |
| 70 | ctx-lh-precuneus | MeanCurv |
| 71 | CC_Mid_Anterior | Itensity normStdDev |
| 72 | ctx-rh-medialorbitofrontal | ThickStd |
| 73 | ctx-lh-precuneus | GausCurv |
| 74 | wm-rh-lateraloccipital | Intensity normMax |
| 75 | ctx-lh-parahippocampal | ThickStd |
| 76 | wm-lh-parahippocampal | Number of Voxels |
| 77 | Right-Lateral-Ventricle | Itensity normStdDev |
| 78 | ctx-rh-medialorbitofrontal | MeanCurv |
| 79 | Left-Amygdala | Volume |
| 80 | ctx-lh-parahippocampal | GrayVol |
| 81 | Right-UnsegmentedWhiteMatter | Intensity normMean |
| 82 | wm-rh-entorhinal | Itensity normStdDev |
| 83 | ctx-rh-pericalcarine | FoldInd |
| 84 | wm-lh-isthmuscingulate | Volume |
| 85 | wm-lh-transversetemporal | Number of Voxels |
| 86 | Left-Hippocampus | Intensity normRange |
| 87 | ctx-lh-paracentral | NumVert |
| 88 | ctx-lh-precuneus | CurvInd |
| 89 | Brain-Stem | Itensity normStdDev |
| 90 | ctx-rh-entorhinal | ThickStd |
| 91 | ctx-rh-inferiortemporal | CurvInd |
| 92 | ctx-rh-lateraloccipital | ThickAvg |
| 93 | ctx-rh-cuneus | GrayVol |
| 94 | ctx-rh-frontalpole | ThickAvg |
| 95 | ctx-rh-superiortemporal | GrayVol |
| 96 | ctx-lh-inferiorparietal | CurvInd |
| 97 | ctx-lh-transversetemporal | GrayVol |
| 98 | wm-lh-middletemporal | Intensity normMax |
| 99 | wm-lh-bankssts | Number of Voxels |
| 100 | ctx-lh-bankssts | MeanCurv |
